# Using an Agent-Based Model to Assess K-12 School Reopenings Under Different COVID-19 Spread Scenarios – United States, School Year 2020/21

**DOI:** 10.1101/2020.10.09.20208876

**Authors:** Timothy C. Germann, Manhong Z. Smith, Lori Dauelsberg, Geoffrey Fairchild, Terece L. Turton, Morgan E. Gorris, Chrysm Watson Ross, James P. Ahrens, Daniel D. Hemphill, Carrie Manore, Sara Y. Del Valle

**Affiliations:** Physics & Chemistry of Materials Group, Los Alamos National Laboratory; Information Systems & Modeling Group, Los Alamos National Laboratory; Center for Nonlinear Studies, Los Alamos National Laboratory; Applied Computer Science, Los Alamos National Laboratory; Computer Science Department, University of New Mexico; National Security Education Center, Los Alamos National Laboratory; Advanced Research in Cyber Systems, Los Alamos National Laboratory

## Abstract

School-age children play a key role in the spread of airborne viruses like influenza due to the prolonged and close contacts they have in school settings. As a result, school closures and other non-pharmaceutical interventions were recommended as the first line of defense in response to the novel coronavirus pandemic (COVID-19). Assessing school reopening scenarios is a priority for states, administrators, parents, and children in order to balance educational disparities and negative population impacts of COVID-19. To address this challenge, we used an agent-based model that simulates communities across the United States including daycares, primary, and secondary schools to quantify the relative health outcomes of reopening schools. We explored different reopening scenarios including remote learning, in-person school, and several hybrid options that stratify the student population into cohorts (also referred to as split cohort) in order to reduce exposure and disease spread. In addition, we assessed the combined impact of reduced in-person attendance in workplaces (e.g., through differing degrees of reliance on telework and/or temporary workplace closings) and school reopening scenarios to quantify the potential impact of additional transmission pathways contributing to COVID-19 spread. Scenarios where split cohorts of students return to school in non-overlapping formats resulted in significant decreases in the clinical attack rate (i.e., the percentage of symptomatic individuals), potentially by as much as 75%. These split cohort scenarios have impacts which are only modestly lesser than the most impactful 100% distance learning scenario. Split cohort scenarios can also significantly avert the number of cases–approximately 60M and 28M–depending on the scenario, at the national scale over the simulated eight-month period. We found the results of our simulations to be highly dependent on the number of workplaces assumed to be open for in-person business, as well as the initial level of COVID-19 incidence within the simulated community. Our results show that reducing the number of students attending school leads to better health outcomes, and the split cohort option enables part-time in-classroom education while substantially reducing risk. The results of this study can support decisions regarding optimal school reopening strategies that at the population level balance education and the negative health outcomes of COVID-19.

**Disclaimer:** This work was sponsored by the United States Centers for Disease Control and Prevention. Los Alamos National Laboratory, an affirmative action/equal opportunity employer, is operated by Triad National Security, LLC, for the National Nuclear Security Administration of the United States Department of Energy under contract # 19FED1916814CKC. Approved for public release: LA-UR-20-27982.

The findings and conclusions in this report are those of the authors and do not necessarily represent the official position of the Centers for Disease Control and Prevention or Los Alamos National Laboratory.

## 1. Introduction

The novel coronavirus disease (COVID-19) was first identified in Wuhan, China in late December 2019 [3] and subsequently spread worldwide. By March 11, 2020, when the World Health Organization (WHO) declared COVID-19 as a global pandemic, there were already close to 120,000 confirmed cases and more than 4,300 deaths worldwide [4]. As of October 8, 2020, there are now over 36 million confirmed cases and over a million deaths worldwide, with over 7.8 million confirmed cases and over 217,000 deaths in the U.S. [5]. The COVID-19 virus spreads primarily through small droplets and aerosols of saliva or discharge from the nose of an infected person [6]. At this time, there are no specific vaccines or large-scale treatments for COVID-19 [6], demonstrating the urgent need for non-pharmaceutical approaches that could reduce its spread.

Public officials have recommended a range of individual- and community-level non-pharmaceutical interventions to slow the spread of COVID-19 and mitigate the impact on people, communities, and healthcare infrastructure [7]. Individual measures include personal protective actions, such as applying proper cough etiquette in daily life, hand hygiene, wearing face coverings/ masks, staying home when sick (also called isolation), or staying home after an exposure to a confirmed case or after residing in/arriving from a community with known widespread transmission (also called quarantine). Community measures may include temporary school closures/dismissals and other social distancing measures such as stay-at-home recommendations, canceling mass gatherings, and minimizing face-to-face contact at workplaces.

As the cases of COVID-19 started to emerge during the early spring of 2020 in the U.S., most of the primary and secondary schools closed for the remainder of the 2019-2020 school year [8]. There has been a significant debate about school closures and reopenings because of the existing educational disparities that have been exacerbated by the pandemic, social isolation, and other unintended consequences such as access to free and subsidized lunches at school. However, there is anecdotal evidence that reopening schools, for the traditional academic year in autumn 2020, in areas experiencing widespread community transmission provide additional transmission pathways between communities that were otherwise mostly isolated. For example, the Cherokee County School District in Georgia reported 108 confirmed cases of COVID-19 within two weeks of schools reopening, and 3 out of the 6 high schools in the district reverted to full remote leaning by the third week [9, 10]. In Mississippi, 71 of the 82 counties reported positive COVID-19 cases within few weeks of schools reopening [11]. Similarly, in Tennessee, over 2,000 children tested positive for COVID-19 within two weeks of schools reopening [12].

Mathematical and computational models of COVID-19 spread provide a platform to examine which modalities of in-person instruction may be feasible during the ongoing COVID-19 pandemic. Several recent studies have begun to quantify the impact of various non-pharmaceutical interventions in combination with different school reopening strategies and have found that reopening schools as normal is likely to increase the number of COVID-19 cases [13, 14]. Other studies have found that closing schools and incorporating social distancing measures in classrooms are effective in reducing the spread of COVID-19 [15]. In addition, hybrid approaches to learning, such as capping the in-person classroom size, may be effective in reducing transmission [14,16] and provide a balance approach between supporting education while limiting the spread of COVID-19.

An additional challenge of simulating the impacts of COVID-19 within a community is also simulating workplace restrictions, which may reduce transmission pathways within the community. We address this gap by combining non-pharmaceutical interventions, school reopening scenarios, and workplace restrictions into an agent-based model, EpiCast, to assess the potential feedbacks on the spread of COVID-19. Using parameters provided by the Centers for Disease Control and Prevention (CDC) to simulate COVID-19 transmission within the U.S., we explored several reopening scenarios including remote learning, in-person school, and several hybrid options that stratify the student population into cohorts in order to reduce exposure and disease spread. The results of this study can support decisions regarding optimal school reopening strategies that balance education and the negative health outcomes of COVID-19.

## 2. Methods

### Model Description

We used an agent-based model, known as Epidemiological Forecasting (EpiCast), originally designed to simulate community-level influenza transmission in the U.S. at the national-scale and adapted it to simulate COVID-19 [17]. The primary modifications for COVID-19 relate to the disease natural history (as described later) since the transmission mechanisms for COVID-19 are similar to that for influenza. The national-scale simulation model consists of 281 million individuals distributed among 65,334 census tracts to closely represent the actual population distribution according to the 2000 U.S. Census data [17]. Each tract is organized into 2,000-person communities resulting in 180,492 model communities. The model combines U.S. Census demographics and worker-flow data to generate daytime and evening contact networks based on potential contacts emerging at daycares, schools, workplaces, households, neighborhoods (∼500 people), and communities (e.g., mall, supermarket) [17]. In each census tract, the synthetic population matches the actual population in several statistical measures including the number of residents and households, the household’s age distribution, the household size and membership distribution, and employment status for working adults. In addition, each workplace is assigned a 3-digit NAICS (North American Industrial Classification System) code based on the proportion of workers in each sector in each county. We used a regional model (∼8.6 million people in the Chicago Metropolitan Statistical Area (MSA)) to explore additional scenarios (given the extensive computational nature of the national model) in order to determine the impact of different assumptions on COVID-19 spread and mitigation strategies.

A new feature of EpiCast, for the purpose of this study, is the ability to capture interactions between teachers and students while in school settings. In previous EpiCast simulation models [17,18], school mixing groups accounted only for transmission between students; teachers and staff were not explicitly included. For the present study, we associate a workplace with NAICS Subsector Code 611 (Educational Services) with each school, and account for mixing between the teachers, staff, and school children. Where necessary, we add additional workplace(s) in a community to achieve an average 14:1 student:(teacher/staff) ratio in each school, based on recent statistics from the National Center for Education Statistics [19]. This is necessary because our community model assumes that elementary and secondary school children attend school in the tract/community in which they reside, not accounting for bussing across Census tract boundaries which the actual employment statistics reflect. Transmission between children in a school mixing group, and between teachers/staff in a workplace mixing group, are unchanged from the original model. For the added mixing, from students to teachers/staff and vice versa, we assume that the individual child-adult contacts are twice the child-child contact rates. Our results were not overly sensitive to this assumption, and we note that the numbers of child-child transmissions are still greater than child-adult transmission due to the much larger number of children in a school. For example, if there are approximately 14 times more children than adults in a school, and approximately two times greater transmission between an individual child and individual adult, the child-child transmission will be about seven times greater than child-adult transmission.

### Epidemiological Parameter Assumptions

In order to simulate COVID-19 transmission within a community, we used parameter assumptions and model-produced epidemiological data from the CDC’s Pandemic Planning Scenarios [20] (Table 1). The disease natural history for COVID-19 was assumed to be as follows: the distribution of latent infection is 1-7 days, the incubation period is 1-8 days, and infectious period is 3-9 days. Furthermore, the proportion of infections which remain asymptomatic are assumed to be 40% and the relative infectiousness of asymptomatic or pre-symptomatic individuals is assumed to be 75%. Self-isolation of symptomatic individuals is assumed to be similar to those used for pandemic influenza studies [21]. Assumptions regarding ideal reduction in contacts due to social distancing, facemasks, and hygiene is shown in Table 2. The “reduced” social distancing scenarios assume a 50% reduction in compliance of preschool and elementary school-age children to account for limited facemask or social distancing measures. Finally, long distance travel is assumed to be reduced due to travel and quarantine restrictions implemented across the nation (Table 2). Each county was initialized and calibrated to match the cumulative case counts during the first two weeks of August 2020 as reported by the New York Times COVID-19 repository [22]. Note that we do not report the number of cases during the calibration phase and thus assume that the simulation starts on August 15, 2020.

**Table 1.** Summary of Key EpiCast model parameters for this study.

| Parameter | Age Group |  |  |  |
| --- | --- | --- | --- | --- |
|  | 0-49 | 50-64 | 65+ | Overall |
| **Symptomatic case hospitalization rate** | 0.017 | 0.045 | 0.074 | 0.034 |
| **Symptomatic case fatality rate** | 0.0005 | 0.002 | 0.013 | 0.004 |
| **Percentage of hospitalized cases requiring treatment in the ICU** | 23.6% | 36.2% | 35.1% | - |
| **Percentage of hospitalized cases requiring $\geq 1$ day of ventilator use** | 11.7% | 21.8% | 21.3% | - |

**Table 2.**
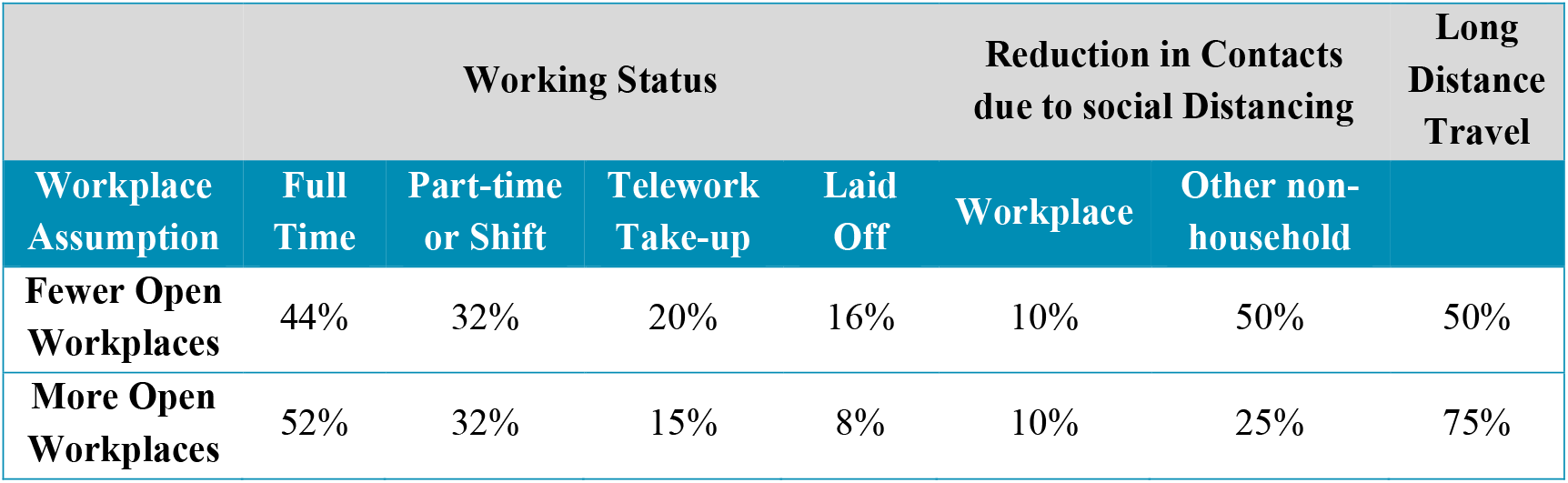
Workforce status & reduction in contacts due to social distancing assumptions.

Assumptions regarding full time, part-time, the number of individuals teleworking, and employees laid off as a result of the current COVID-19 situation are shown in Table 2. Some of these percentages were chosen based on discussions with subject matter experts from the State of New Mexico. Furthermore, the percentage of individuals teleworking are based on two surveys of the labor market near the beginning of the COVID-19 pandemic in the U.S. from the Bureau of Labor Statistics (BLS) [23]. The ability to telework for each 3-digit NAICS sector also comes from the BLS survey and is shown in Table 3. The model assumptions on working in the workplace versus working from home or being laid off were based on the values in both Table 2 and Table 3.

**Table 3.**
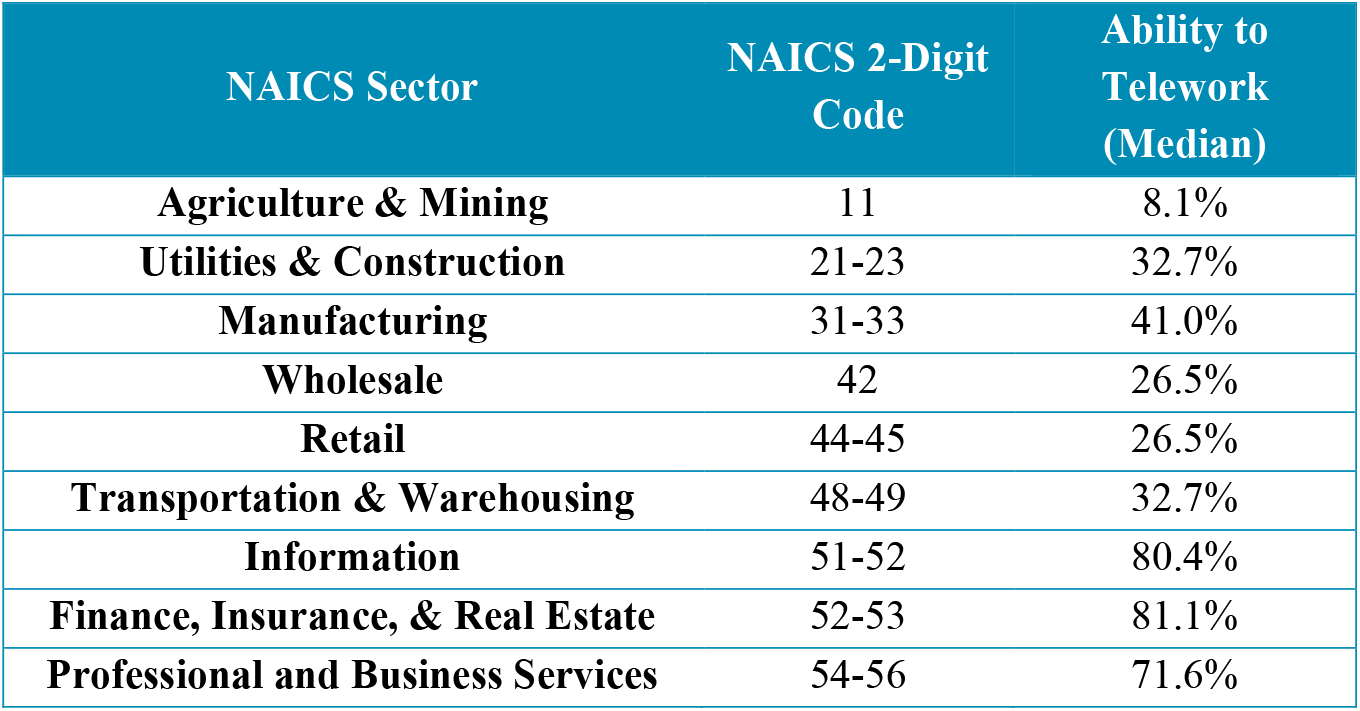
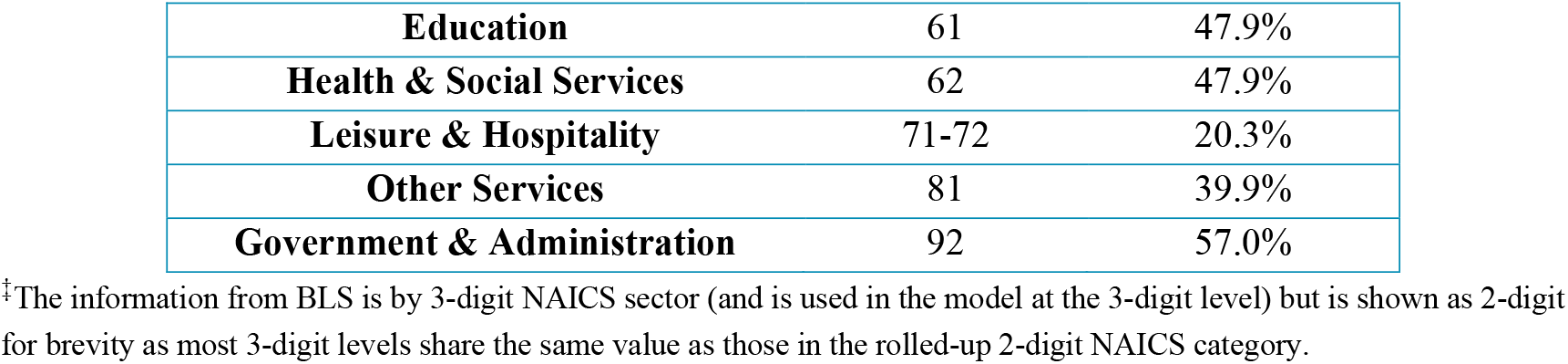
Ability to Telework by NAICS 2-Digit Sector.^‡^

### Workplace Modeling Assumptions

Per the phase guidelines released in Opening Up America Again [24], we modeled two scenarios: “Fewer Open Workplaces,” similar to Phase 2 of Opening Up America Again, and “More Open Workplaces”, similar to Phase 3. These two scenarios describe different levels of in-person workplace assumptions (Figure 1, Tables 2-3). Specifically, Fewer Open Workplaces encourages telework whenever possible and feasible with business operations as well as limited onsite operations for a small set of businesses. More Open Workplaces assumes staffing of additional worksites with an expanded number of onsite workers. An example is a retail business may be open to 25% customer capacity as per Phase 2 recommendations and the NAICS industry percentage of employees working onsite is 50% (in order to accommodate the workers necessary for the operation of the business) for Fewer Open Workplaces. For More Open Workplaces, this business may have the opportunity to have a 50% customer capacity and the percentage of employees needed would increase to 75%. For a comparison across intervention approaches, we also use a Pre-pandemic Behavior scenario, which assumes that all businesses are open with no capacity or social distancing restrictions.

**Figure 1.**
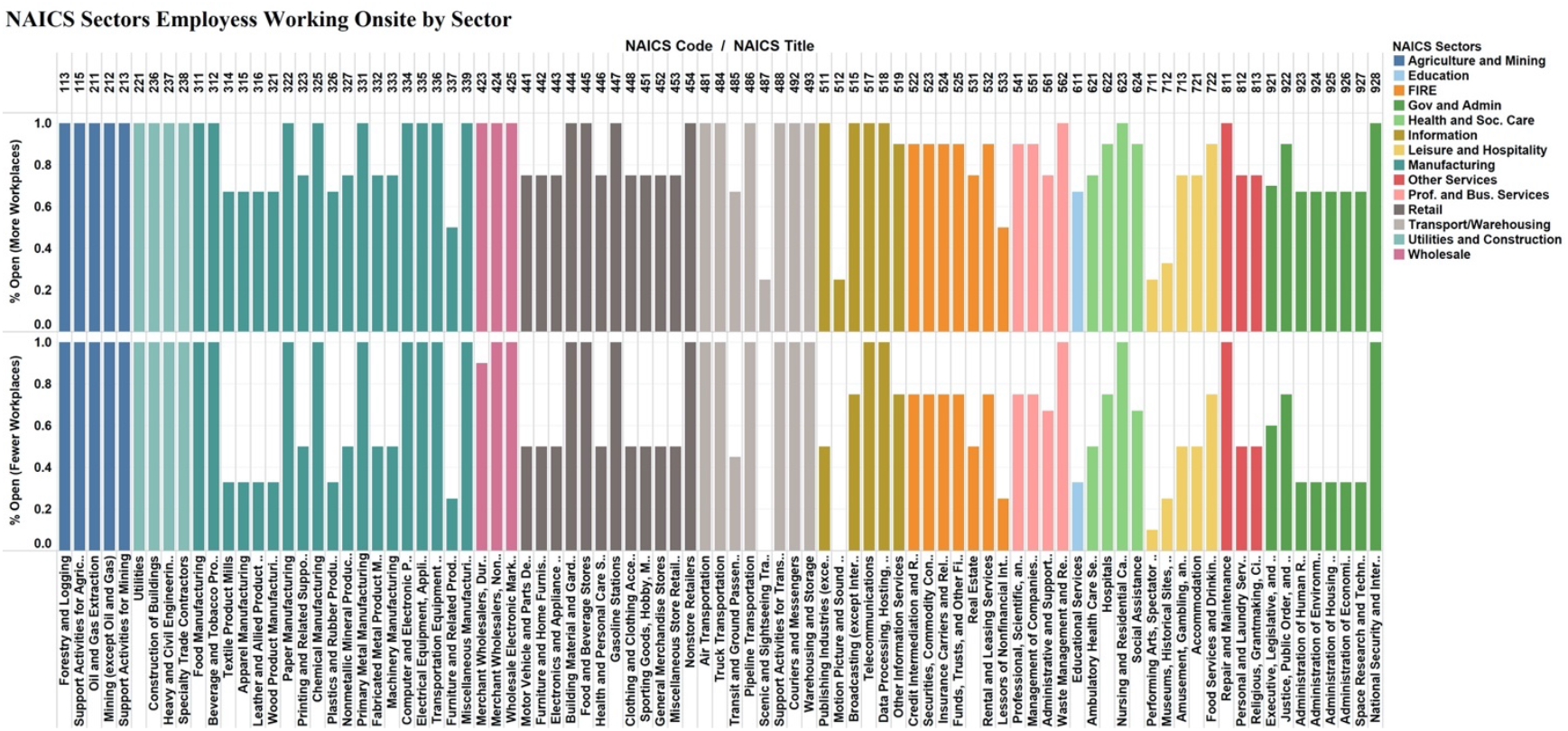
NAICS sectors assumptions for Fewer Open Workplaces and More Open Workplaces.

### School Scenarios

We explored the impact of various school reopening scenarios as described in Table 4. These scenarios range from 100% distance learning to 100% onsite learning (Baseline), as well as partial onsite learning with alternating days or weeks. We also explored 80% in-person enrollment due to recent surveys, which suggest that at least 20% of parents may not send their children back to school [25]. For the regional model, we assume that the 2020-2021 school year for the Chicago Public Schools begins on September 8^th^, 2020. The national scale model assumes a different start date for the 2020-2021 school year ranging from August 3^rd^, 2020 (Arizona) to September 16^th^, 2020 (New York) based on publicly available information from school districts.

**Table 4.** Descriptions of school reopening, baseline, and pre-pandemic scenarios.§

| Scenario Name | Scenario Code | Scenario Description |
| --- | --- | --- |
| Pre-Pandemic Behavior | Pre-Pandemic Behavior | No mitigations, all businesses completely open (i.e., 100% enrollment with no social distancing in place). |
| Baseline | Baseline | All students physically in school with some social distancing (i.e., 100% enrollment). |
| 80% Onsite Learning with **Reduced** Social Distancing‡ | 80%_OL_LessSD | All enrolled* students physically in school. |
| 80% Onsite Learning with **Ideal** Social Distancing† | 80%_OL_SD | All enrolled* students physically in school. |
| 80% Partial Onsite Learning – **Alternating Week** with **Reduced** Social Distancing‡ | 40%_POL_LessSD_Week | Two non-overlapping cohorts of students – 40% of the students attend one week and the other 40% attend the next week. |
| 80% Partial Onsite Learning – **Alternating Days** with **Reduced** Social Distancing‡ | 40%_POL_LessSD_2Day | Two non-overlapping cohorts of students – 40% of the students attend for two days/week (Mon/Tue) and the other 40% attend for two days (Thu/Fri). Wednesday off for disinfection. |
| 80% Partial Onsite Learning – **Alternating Weeks** with **Ideal** Social Distancing† | 40%_POL_SD_Week | Two non-overlapping cohorts of students – 40% of the students attend one week and the other 40% attend the next week. |
| 80% Partial Onsite Learning – **Alternating Days** with **Ideal** Social Distancing† | 40%_POL_SD_2Days | Two non-overlapping cohorts of students – 40% of the students attend for two days/week (Mon/Tue) and the other 40% attend for two days (Thu/Fri). Wednesday off for disinfection. |
| 100% Distance Learning | Offsite | No students physically in school. |

§Note that given the similar results between all the “Less SD” scenarios in the regional simulations, we did not run the Less SD for the nationwide scenarios.
\*Note that 80% in-person enrollment was used due to recent surveys that suggest that at least 20% of parents may not send their children back to school [25].
† *Ideal social distancing* assumes 50% reduction in contacts due to students staying 6 ft from other people, increased hygiene, and masks/face coverings.
‡ *Reduced social distancing* assumes 25% reduction in contacts of preschool and elementary school-age children to account for limited facemask use or limited social distancing measures.

## 3. Results

### Overall Regional and National Impacts

The impacts of school reopening for the Chicago MSA region are summarized in Figure 2 and Tables A-1 and A-2 in the Appendix. Figure 2 shows the epidemic curves for nine school reopening scenarios under Fewer Open Workplaces and More Open Workplaces for the Chicago MSA region aggregated over the simulated eight-month period (15 August 2020 through 11 April 2021). Figure 3 shows the national simulation results of six scenarios under Fewer Open Workplaces and More Open Workplaces for the nation. The results show similar trends for the Baseline, 80% in-person learning, 40% 2-day and alternating week, and offsite school scenarios with Fewer Open Workplaces and More Open Workplaces assumptions for both regional and national simulations.

**Figure 2.**
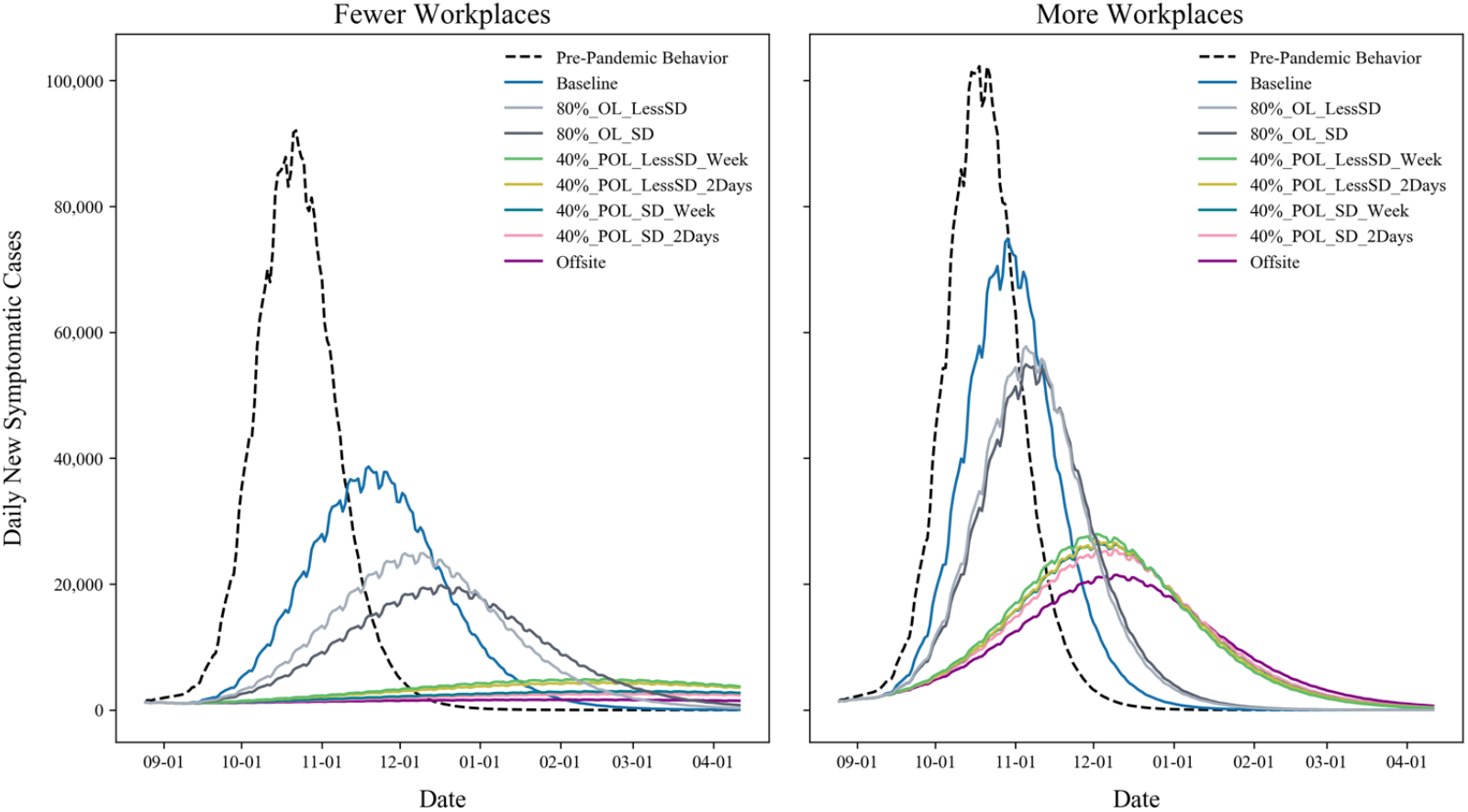
Chicago MSA results from the EpiCast model for various school opening scenarios under Fewer Open Workplaces and More Open Workplaces.

**Figure 3.**
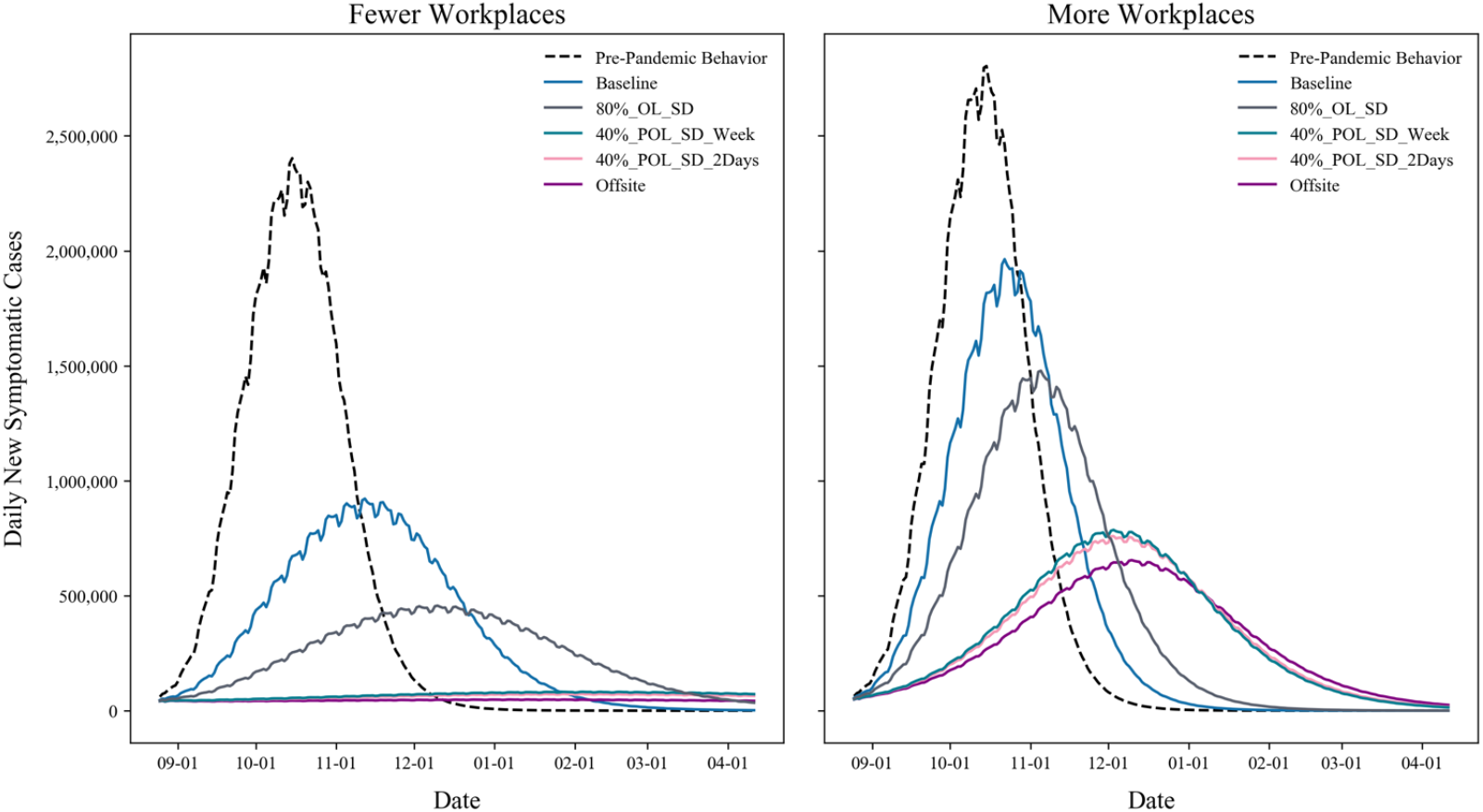
National results from the EpiCast model for various school opening scenarios under Fewer Open Workplaces and More Open Workplaces.

All the partial onsite learning scenarios delay the epidemic peak and flatten the curve for Fewer Open Workplaces, which is consistent with previous studies on school closures [13-16] (Figure 2, 3). However, for More Open Workplaces, the peak for most scenarios is spread around three weeks regardless of school reopening scenario and the impact of hybrid school reopenings is reduced. Additionally, the reduced social distancing scenario (analyzed only the Chicago MSA region), which accounts for limited compliance in facemask usage and social distancing measures for children in K-8 (i.e., kindergarten through grade 8^th^), has a slight but significant decrease in the clinical attack rate (CAR) (i.e., the percentage of symptomatic individuals) over the simulated eight-month period. That is, for the Fewer Open Workplaces scenario, the CAR is reduced from 26.3% for the 80% less social distancing scenario to 23.8% for the ideal social distancing scenario. Similarly, the CAR is reduced for the 40% 2-day split cohort scenario from 8.8% to 5.8% for the Fewer Open Workplaces scenarios depending on the ideal or reduced social distancing assumptions, respectively. These results show that reducing the number of students attending in-person education as well as splitting the student population into cohorts, can reduce the potential negative impacts of COVID-19 spread.

The results show heterogeneity in the impacts across the U.S. Figures 4-5 show cumulative cases per 100K population at the county level for EpiCast simulated results for the 40% split cohort scenario attending 2 days a week and the 80% onsite school scenario. Note that the cumulative number of cases include both symptomatic and asymptomatic individuals aggregated over eight-months.

**Figure 4.**
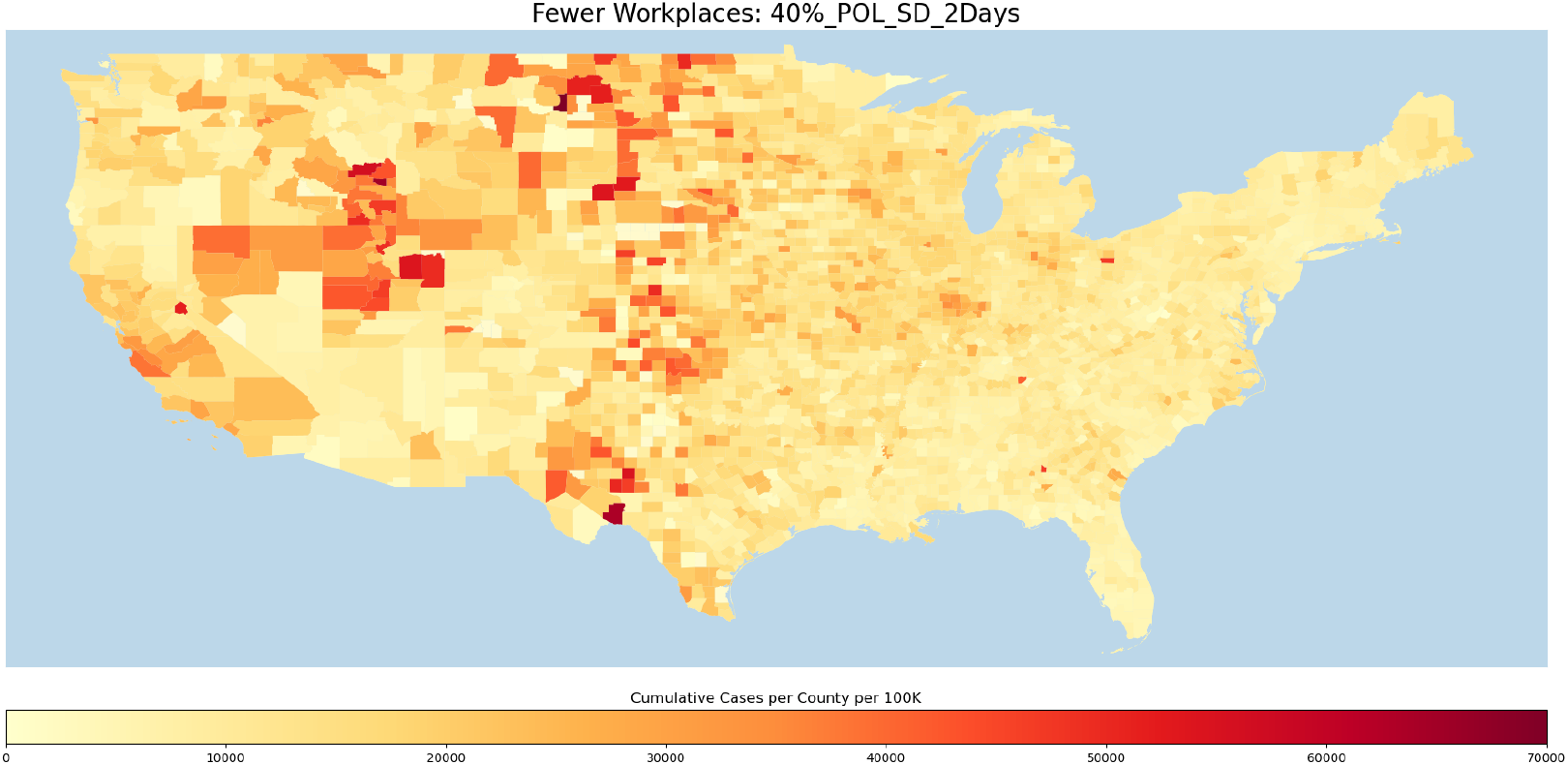
Cumulative cases per county per 100K for EpiCast simulated results for two non-overlapping cohorts of 40% of students attending school 2-days a week.

**Figure 5.**
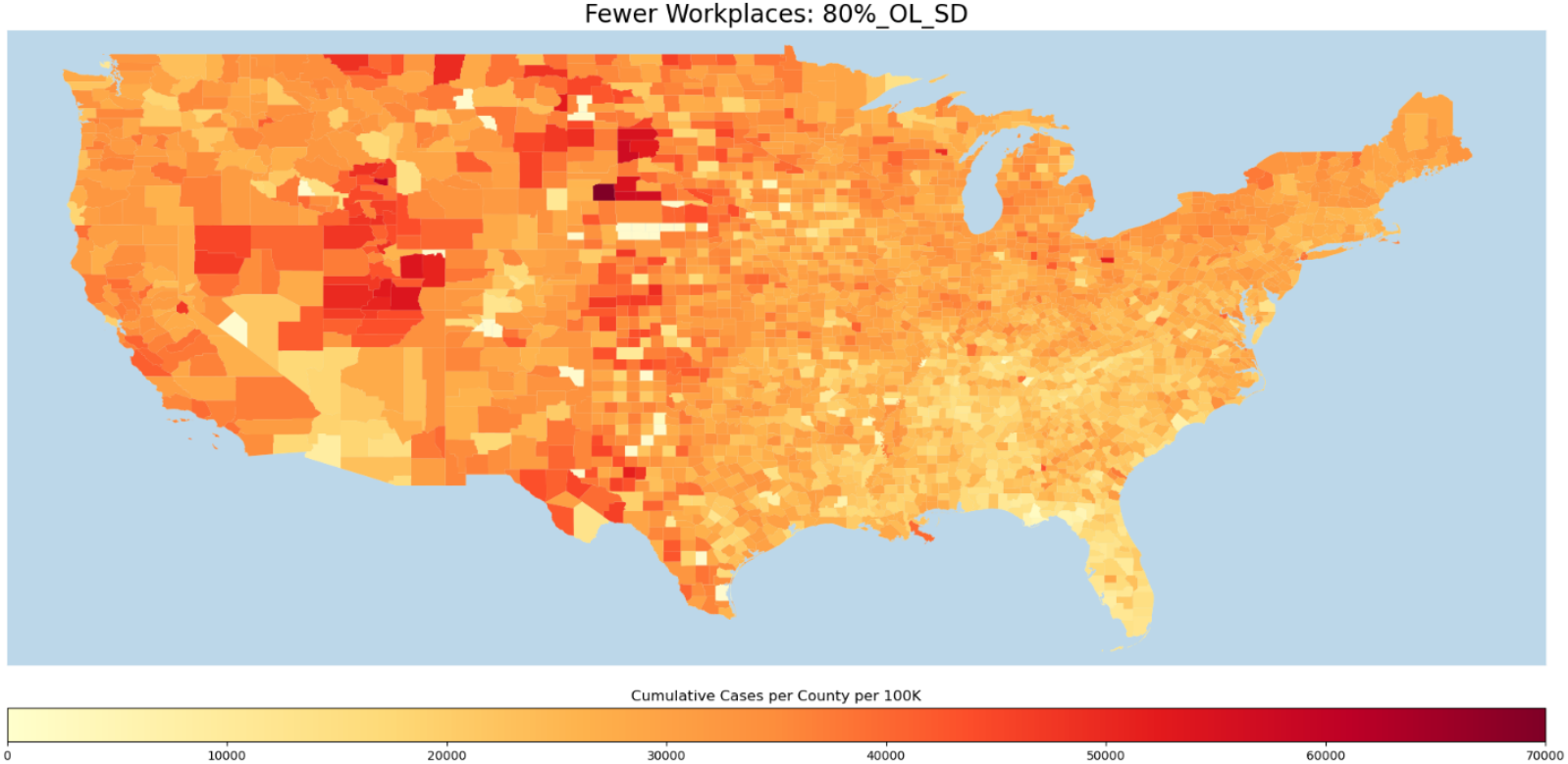
Cumulative cases per county per 100K for EpiCast simulated results for 80% of students attending school full time.

Tables 5-6 show key results from the model aggregated for the nation. Table 5 shows the total number of cases, deaths, and hospitalizations for each scenario for the full eight-month simulation period and for the four weeks around the peak of the epidemic. Table 6 shows the peak incidence and prevalence as well as the time to peak and the total CAR for each scenario. Note that the cumulative number of cases includes both symptomatic and asymptomatic individuals as simulated by EpiCast. Similar impacts as the Chicago MSA model (see Appendix) are observed at the national level but the overall attack rate is lower for all scenarios. Specifically, the scenarios with the lowest attack rate include the 100% offsite, 40% 2-day and alternating weeks school scenarios (i.e., Fewer Open Workplaces: 4.1%, 5.6%, and 6.1%, respectively). As noted earlier, the impacts of COVID-19 spread is lower for the Fewer Open Workplaces compared to the More Open Workplaces for all scenarios.

**Table 5.** Summary of key EpiCast results for the Nation – Part 1

| Workplace Assumptions | Scenario Name | During Peak 4 Weeks |  |  | August 15, 2020 to April 11, 2021 |  |  |
| --- | --- | --- | --- | --- | --- | --- | --- |
|  |  | Cases | Hospitalized | Deaths | Case | Hospitalized | Deaths |
| Fewer Open Workplaces | Pre-Pandemic Behavior | 59,664,577 | 1,798,188 | 107,322 | 110,244,127 | 3,370,360 | 230,451 |
|  | Baseline | 24,323,551 | 685,746 | 38,649 | 75,049,776 | 2,132,798 | 128,292 |
|  | 80%_OL_SD | 12,346,146 | 354,878 | 20,900 | 55,178,391 | 1,588,821 | 95,848 |
|  | 40%_POL_SD_Week | 2,263,045 | 67,090 | 4,108 | 15,922,257 | 466,195 | 27,874 |
|  | 40%_POL_SD_2Days | 1,997,647 | 59,056 | 3,624 | 14,457,662 | 424,601 | 25,474 |
|  | Offsite | 1,336,844 | 39,827 | 2,484 | 10,665,240 | 316,245 | 19,169 |
| More Open Workplaces | Pre-Pandemic Behavior | 68,242,756 | 2,064,544 | 120,162 | 116,608,169 | 3,584,053 | 242,236 |
|  | Baseline | 49,681,358 | 1,470,601 | 84,679 | 102,532,010 | 3,071,051 | 198,517 |
|  | 80%_OL_SD | 38,469,699 | 1,156,296 | 69,342 | 93,355,312 | 2,830,004 | 184,520 |
|  | 40%_POL_SD_Week | 21,206,204 | 657,099 | 42,085 | 75,101,132 | 2,331,432 | 154,298 |
|  | 40%_POL_SD_2Days | 20,479,987 | 636,866 | 41,009 | 73,871,330 | 2,296,792 | 152,097 |
|  | Offsite | 17,756,292 | 556,366 | 36,073 | 68,375,029 | 2,139,919 | 142,522 |

**Table 6.** Summary of key EpiCast results for the Nation – Part 2

| Workplace Assumptions | Scenario Name | Peak Incidence (Cases per 1000) | Time to Peak Incidence (days) | Peak Prevalence (Cases per 1000) | Time to Peak Prevalence (days) | Clinical Attack Rate (%) |
| --- | --- | --- | --- | --- | --- | --- |
| Fewer Open Workplaces | Pre-Pandemic Behavior | 2,402,432 | 62 | 51 | 66 | 42.4 |
|  | Baseline | 922,097 | 90 | 19 | 94 | 28.9 |
|  | 80%_OL_SD | 456,896 | 118 | 10 | 122 | 21.2 |
|  | 40%_POL_SD_Week | 82,209 | 174 | 2 | 178 | 6.1 |
|  | 40%_POL_SD_2Days | 72,686 | 174 | 2 | 178 | 5.6 |
|  | Offsite | 50,110 | 8 | 1 | 157 | 4.1 |
| More Open Workplaces | Pre-Pandemic Behavior | 2,803,605 | 62 | 59 | 65 | 44.8 |
|  | Baseline | 1,963,443 | 69 | 41 | 73 | 39.4 |
|  | 80%_OL_SD | 1,478,282 | 83 | 31 | 87 | 35.9 |
|  | 40%_POL_SD_Week | 786,216 | 111 | 17 | 115 | 28.9 |
|  | 40%_POL_SD_2Days | 758,840 | 111 | 16 | 115 | 28.4 |
|  | Offsite | 654,784 | 118 | 14 | 122 | 26.3 |

### Source of Infection

Identifying the source of infection can help develop targeted mitigations to reduce the potential spread of viruses. Figure 6 shows the source of infection for all the national-level scenarios for Fewer Open and More Open Workplaces for all contact settings within the simulation. Our results show that the majority of cases are generated at home, followed by neighborhood/community settings. The percentage of infection generated in schools and workplaces is correlated with the level of schools/workplaces open. Note that additional infections generated at workplaces are captured under neighborhood/community due to the fact that EpiCast does not explicitly account for customer interactions with workers at workplaces/workgroups. Workgroups only account for infections generated from employee to employee. Figure 7 shows the aggregated source of infection for daycares, playgroups, and schools for all the national-level scenarios for Fewer Open and More Open Workplaces. Note that the majority of school-related infections are generated from student-student interactions due to the prolonged close contacts in these settings. This finding supports strategies that reduce the number of students attending in-person education.

**Figure 6.**
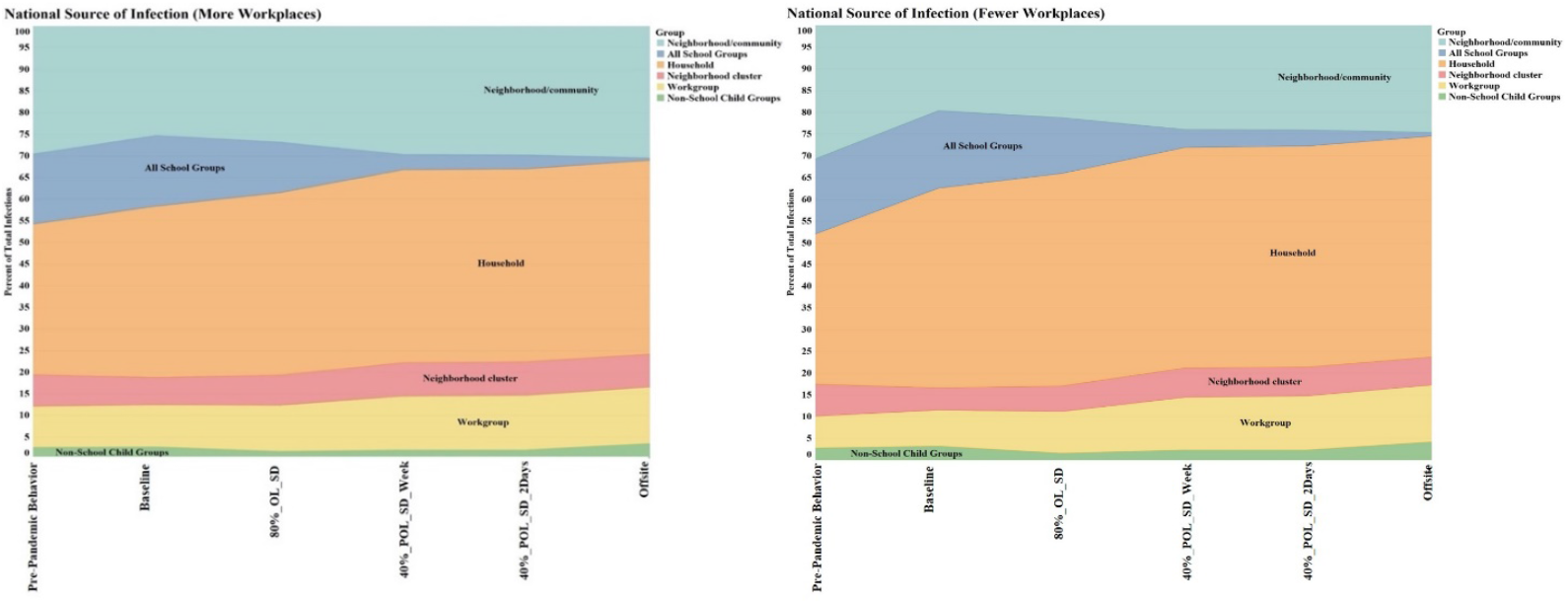
Source of infection for each of the national-level scenarios for Fewer Open and More Open Workplaces. Note that about 30% of the infections are generated at home, 20% at neighborhood/community settings, 15% at schools, and 10% at workplaces.

**Figure 7.**
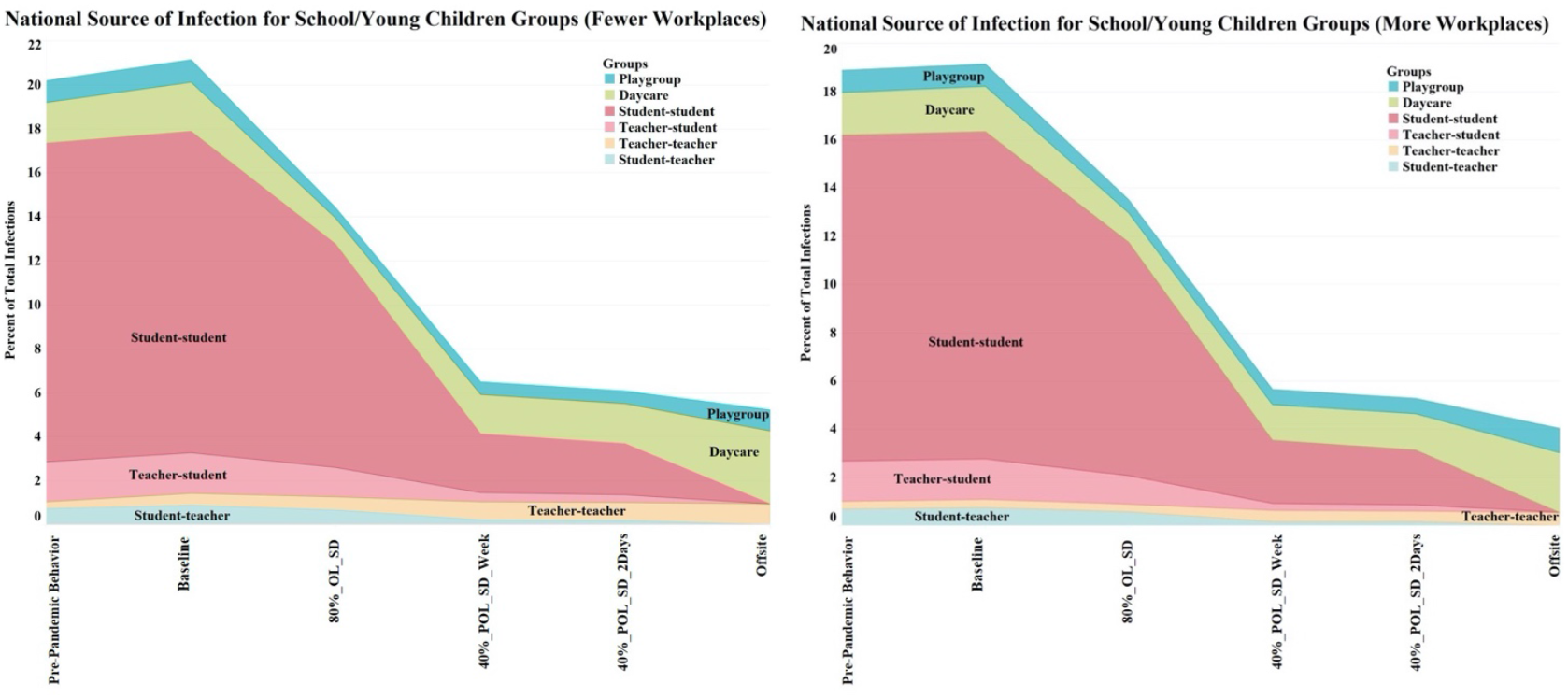
School-related source of infection breakdown including playgroup, daycare, student-student, teacher-student, student-student. The largest contribution is generated from the student-student interactions.

### Cases, Deaths, & Hospitalizations Averted

Non-pharmaceutical interventions are effective in averting the potential cases, deaths, and hospitalizations that would have otherwise resulted without the implementations of these public health strategies. We estimated the number of cases, deaths, and hospitalizations that may be averted by comparing each of the mitigation scenarios against the Baseline scenario. Figure 8 shows the cases *averted* and delay to peak incidence for Fewer Open and More Open Workplaces for all the national-level scenarios. The results show that alternating school cohort scenarios can significantly avert the total number of cases by approximately 60M and 28M for the Fewer Open Workplaces and More Open Workplaces, respectively. The results are consistent with previous studies [18, 26-28] that have shown that non-pharmaceutical interventions can delay the peak of an outbreak (i.e., flatten the curve) and reduce the total number of cases over the same time frame. Furthermore, the offsite scenario provides the largest benefit by averting the greatest number of cases followed by the 40% scenarios. Notably, the 100% distance learning scenario averts nearly 5 million more cases and results in almost twice as long time-to-peak interval compared to the split cohort scenarios. These results demonstrate the positive impacts of non-pharmaceutical interventions in reducing disease burden and flattening the curve to allow for healthcare services not to be overwhelmed. Figure 9 shows deaths and hospitalizations *averted* for Fewer Open and More Open Workplaces for all the national-level scenarios. The results show a significant reduction in deaths and hospitalizations for the 40% 2-day/alternating week and offsite scenarios under Fewer Open Workplaces assumptions. Specifically, over 1.6M hospitalizations and 100K deaths may be averted for all 40% and offsite scenarios under the Fewer Open Workplaces assumptions.

**Figure 8.**
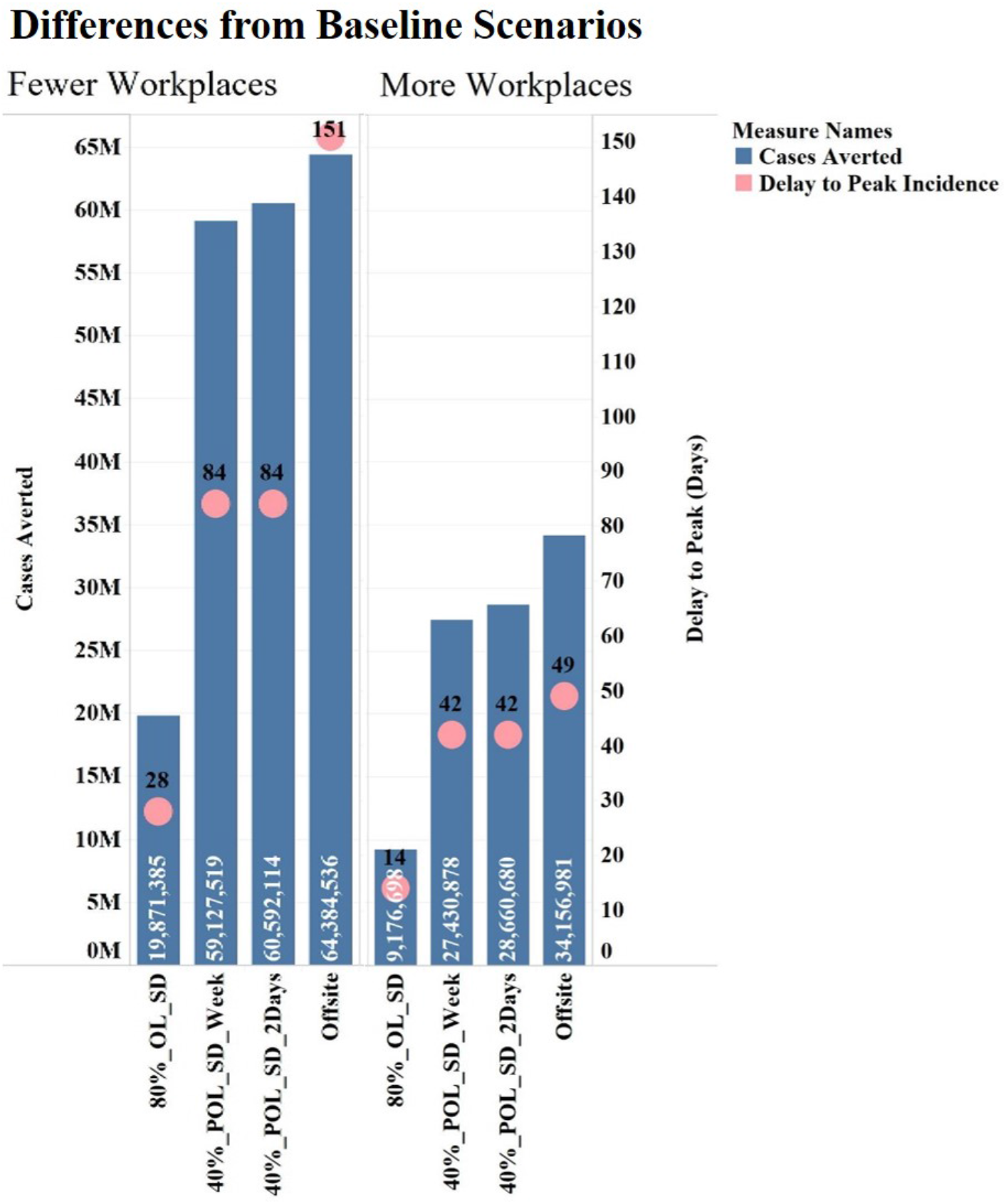
Cases averted and delay to peak incidence in days for all national-level scenarios. Fewer people physically at work and more social distancing along with hybrid school scenarios avert the most cases and delay the peak.

**Figure 9.**
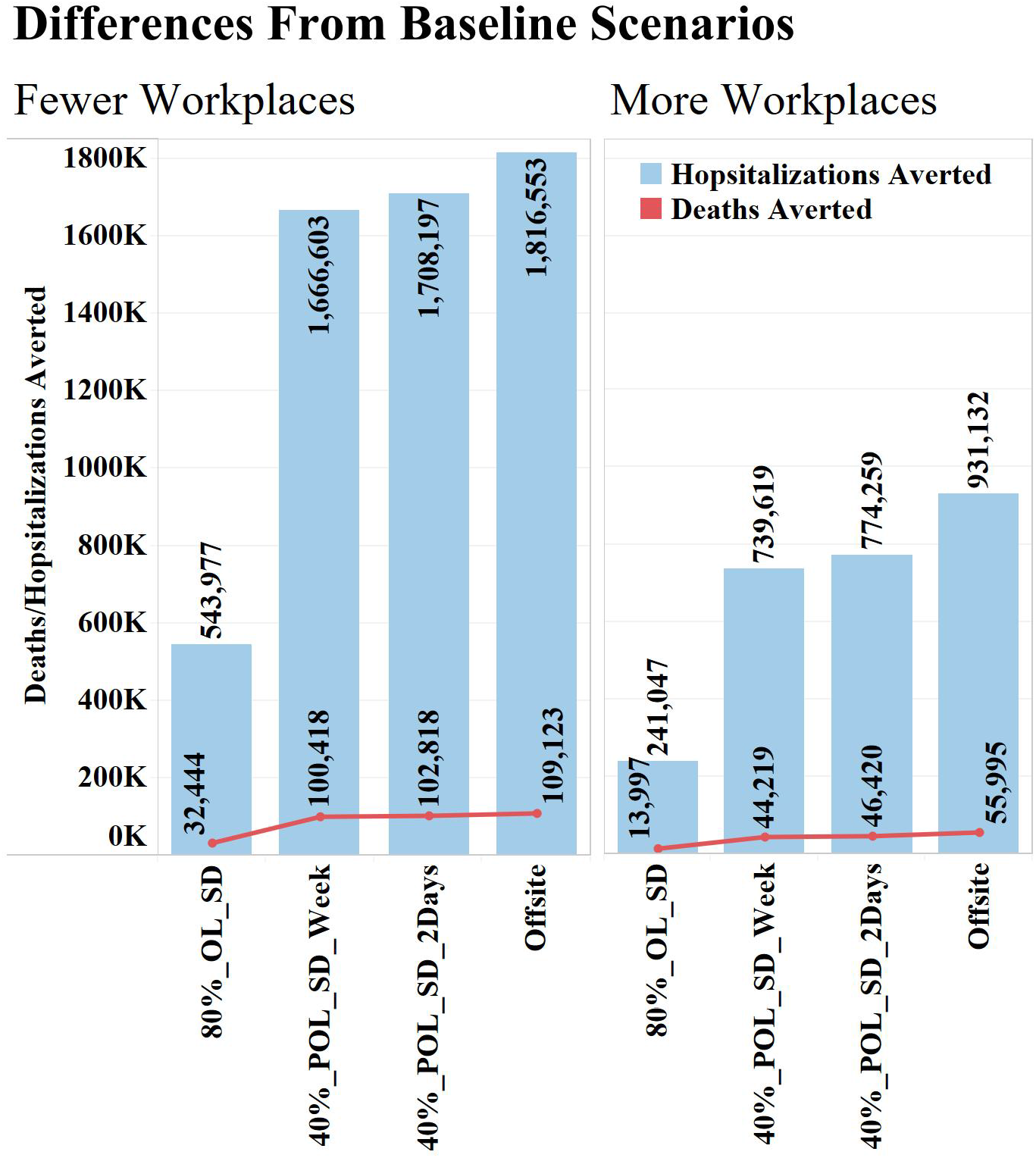
Deaths and hospitalizations averted for all national-level scenarios. There is a significant reduction in deaths and hospitalizations for the 40% 2-day/alternating week and offsite scenarios under Fewer Open Workplaces assumptions

### Impacts by Age

We have observed significant variation in the distribution of cases, hospitalization, and deaths by age throughout the pandemic. Notably, during the early stages of the pandemic, older adults were most affected but the age distribution has changed as the pandemic has progressed [29].Given the demographic granularity of EpiCast, we stratified the total cases, hospitalizations, and ICU beds by age group for all national-level scenarios for Fewer Open Workplaces (Figure 10) and More Open Workplaces (see Appendix and Supplemental Files SF1-SF3). Our results show that the highest number of cases for most scenarios is generated by the 5-18 school-aged group followed by the 30-64 age group. However, adults 30-64 years old make up the largest number of hospitalizations and ICU bed usage for all scenarios. It is worth noting that the age distribution of cases for our simulated scenarios looks different than current COVID-19 age distribution for the nation [30]. While the majority of the cases currently reported were during periods when schools were closed and many children were in isolation, we suspect that as schools open and more children are exposed, the case rates for younger populations may increase. Additionally, there may be other factors contributing to these discrepancies including underreporting due to asymptomatic infections, lack of widespread testing [31], initial parameter estimates, and contact patterns assumptions. Specifically, recent evidence suggests that children are more likely to be asymptomatic [32-34] resulting in biased estimates for this population; however, states that have implemented widespread testing, have reported different age distributions [35]. In addition, the initial planning scenarios and subsequent age breakdowns [20] reflected the hospitalization and case fatality age distributions, not the case distribution of infections, which were closer to the relative population of each age group. Finally, our simulated results used contact patterns based on historical estimates for influenza studies due to lack of contact patterns needed to parameterize our model. However, as more evidence becomes available, we will update our assumptions in order to better assess the impacts of COVID-19 by age.

**Figure 10.**
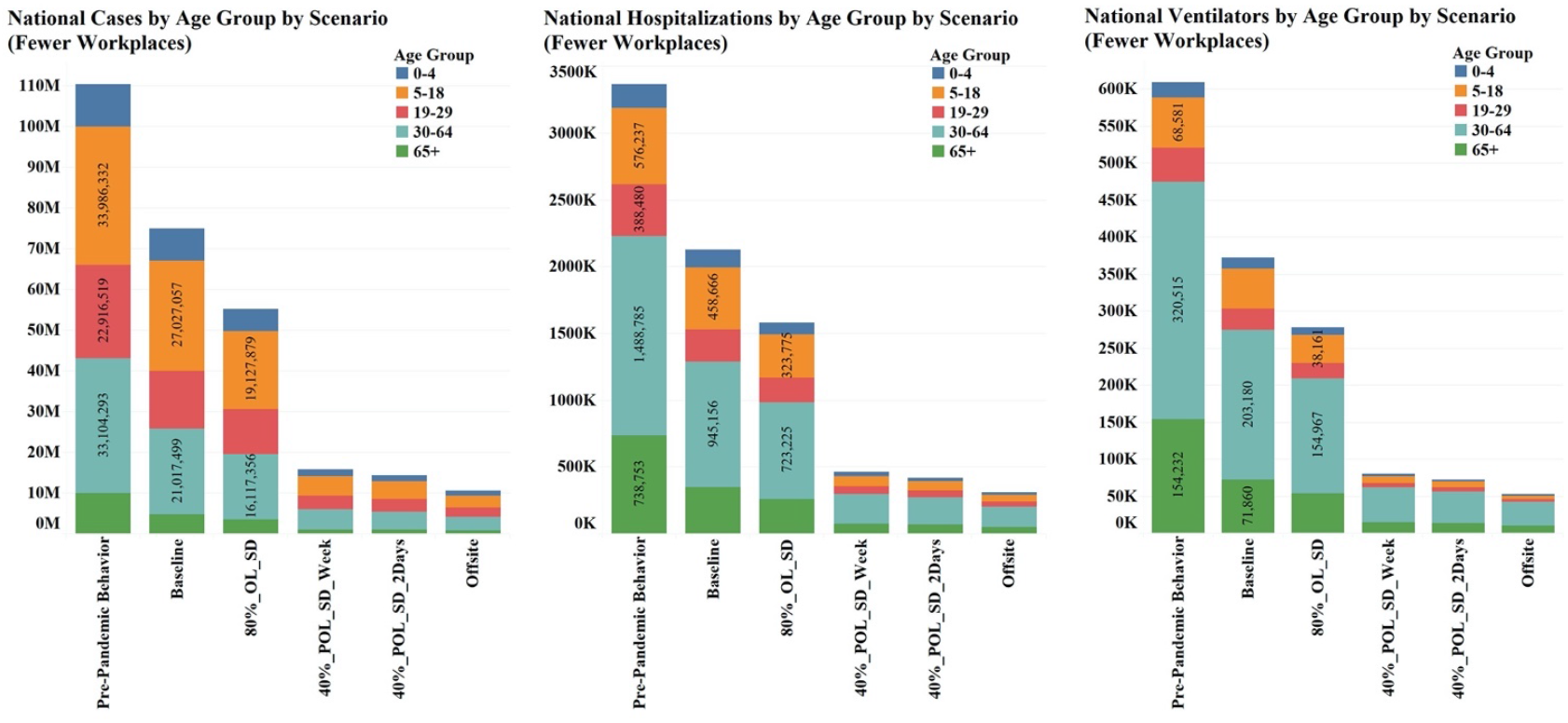
Total cases, hospitalizations, and ICU beds by age group for all the national-level scenarios for Fewer Open Workplaces. Social distancing combined with hybrid school scenarios result in the lowest number of cases.

### State-level Comparisons

Spatial heterogeneity has been evident during the COVID-19 pandemic due to local demographics and public health orders implemented across the states. We selected 12 representative states (at least one state per U.S. Department of Health & Human Services Regions) in order to show the spatial differentiation for all the scenarios. Figures 11-12 show normalized epidemic time series and peak day comparisons for 12 states for the baseline and offsite scenarios (additional figures are included in the Appendix). For the baseline with Fewer Open Workplaces, the peak date ranges from October 21^st^, 2020 (California) to December 10^th^, 2020 (Maine). The peak is dependent on the current transmission levels for each of these states (i.e., states with higher transmission peak first and states with lower transmission peak later). Note that the offsite scenarios result in flat epidemic curves.

**Figure 11.**
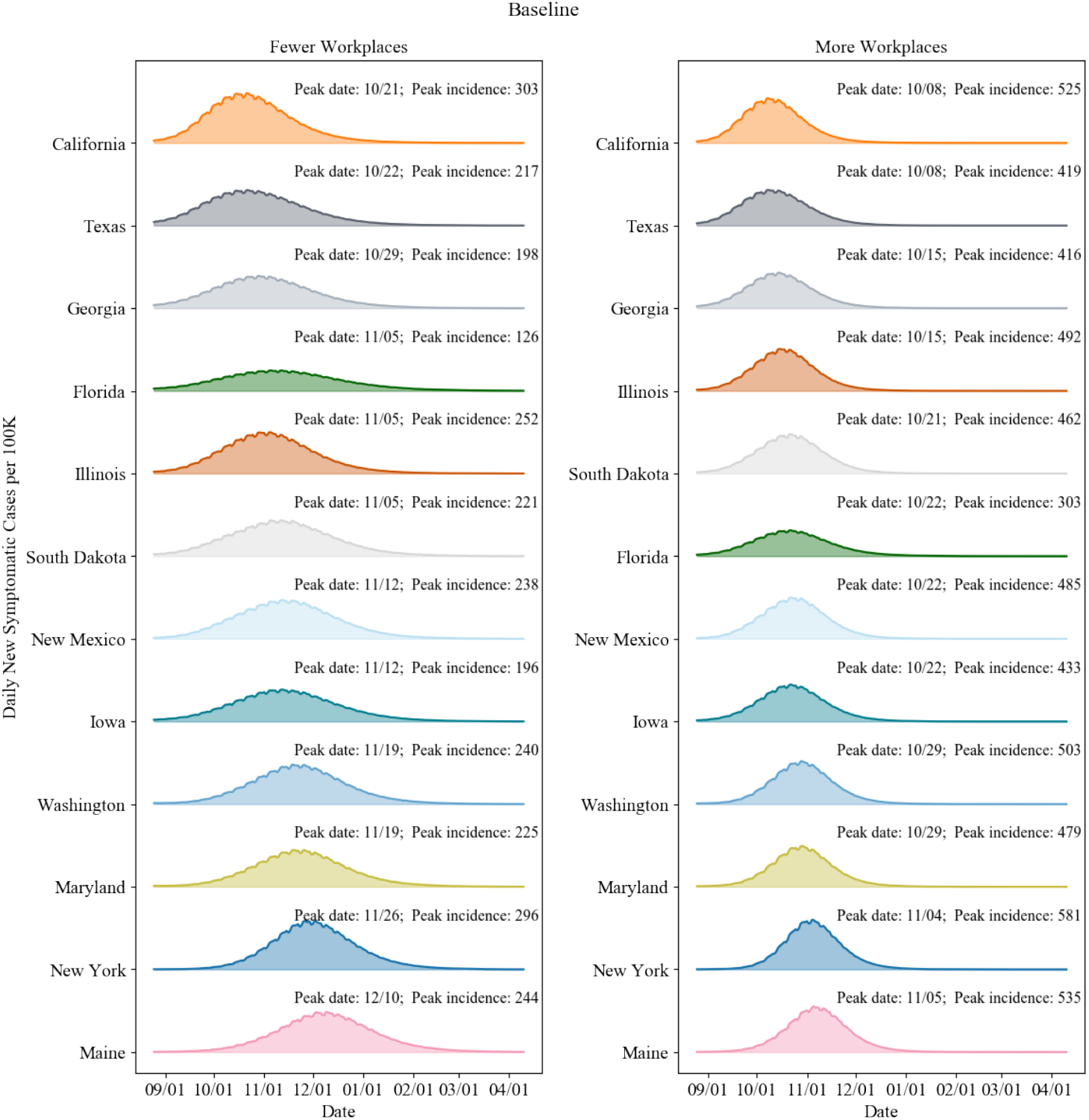
Baseline epidemic curves and peak dates for 12 representative states. (States are sorted by peak date for both scenarios.)

**Figure 12.**
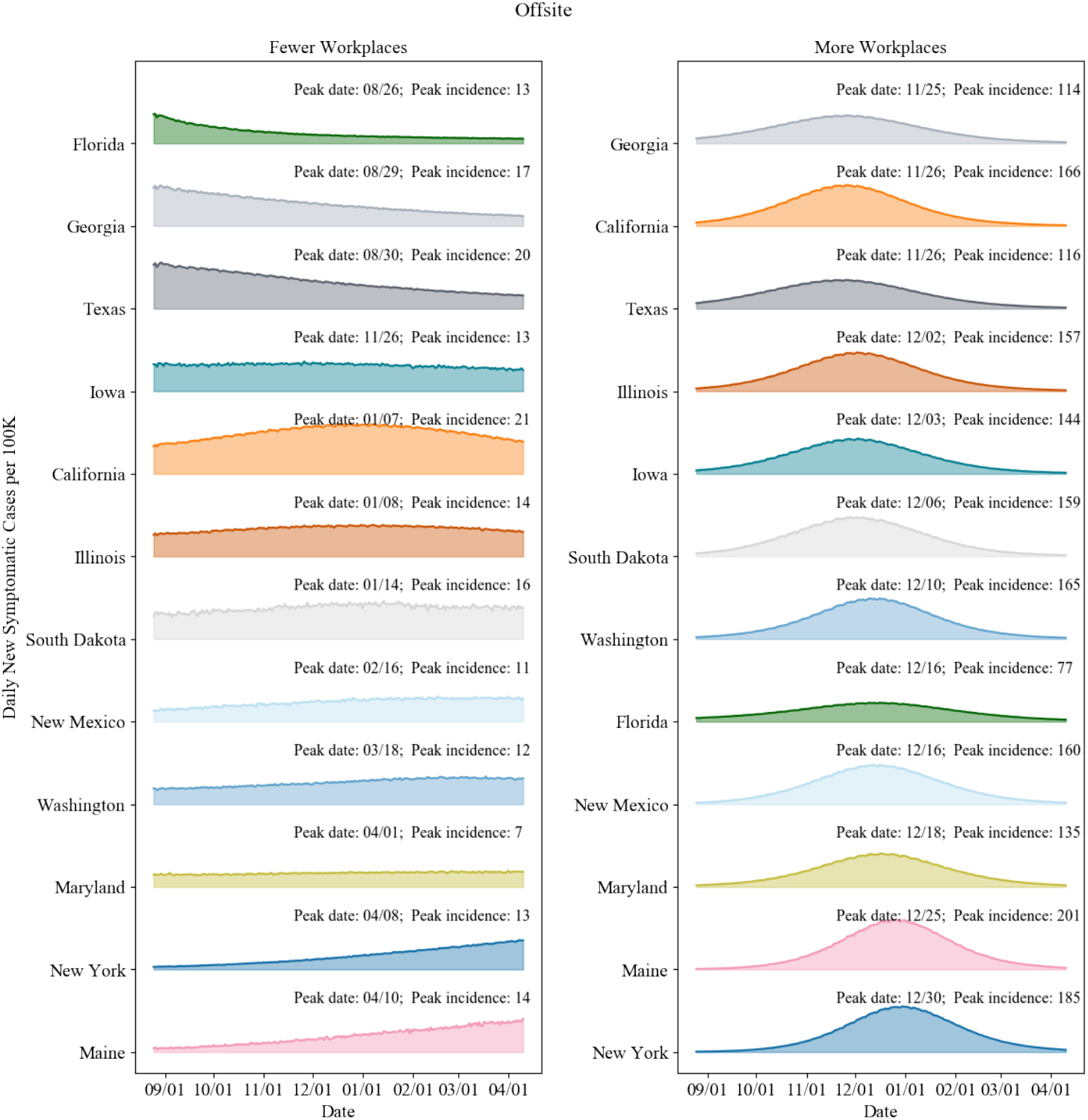
Offsite epidemic curves and peak dates for 12 representative states. (States are sorted by peak date for both scenarios.)

Figure 13 shows epidemic time series and peak day comparisons for 12 states for the 40% 2-day split cohort scenario for both Fewer Open Workplaces and More Open Workplaces. As mentioned earlier, these intervention strategies flatten the curve subsequently delaying the peak. However, the benefits of the cohort scenarios are reduced when more workplaces are open, increasing the transmission paths for the entire population. We note great variability in the impacts and dynamics across the 12 states, especially for the More Open Workplaces scenarios.

**Figure 13.**
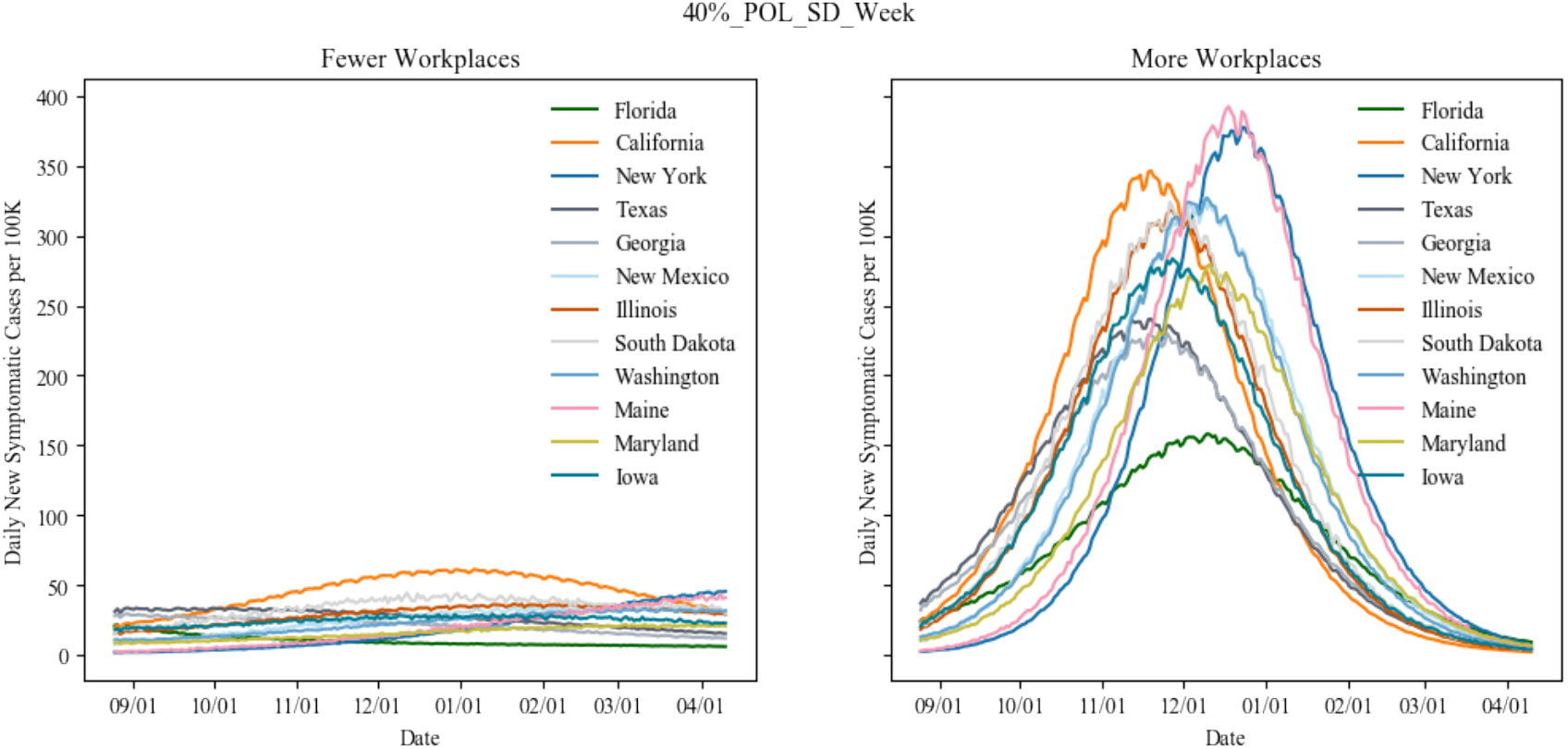
40% onsite alternating week epidemic curves for 12 representative states

## 4. Discussion and Conclusions

Non-pharmaceutical interventions such as school closures and social distancing have been implemented globally to mitigate the spread of COVID-19. Given the start of the new school year, there is a need to assess how to best resume school activities while reducing the risk of increased transmission. We used an agent-based simulation to assess the impact of several school reopening scenarios in combination with community level transmission that accounts for workplace in-person restrictions.

Our results suggest that reducing the number of students by 20% (consistent with the percentage of parents who will likely keep children out of school during the school year 2020/21 [25]) reduces the CAR by at least 5% compared to the ∼100% enrolment, which would be expected in pre-pandemic period. Scenarios where split cohorts of 40% of students return to school in non-overlapping formats may result in more significant decreases in the CAR, potentially by as much as 75%. The split cohort scenarios have impacts which are modestly lesser than the 100% offsite or distance learning scenario. However, the 100% distance learning scenario averts nearly 5 million more cases and results in almost twice as long time-to-peak interval compared to the split cohort scenario. Alternating school cohort scenarios can also significantly avert the total number of cases by approximately 60M and 28M for the Fewer Open Workplaces and More Open Workplaces, respectively. These split cohort scenarios assume appropriate non-pharmaceutical interventions such as social distancing and wearing facemasks at school. Our results indicate that implementing smaller classroom sizes and cohorts of students with breaks between in-person school attendance (e.g., two days on, three days off) can have a major impact on the spread of COVID-19 both in terms of total cases and timing of the peak of a given outbreak wave.

Increasing the number of in-person workplaces (i.e., from Fewer Open to More Open Workplaces) increases the overall CAR for all scenarios. For the alternating school cohort scenarios, there is a nearly five-fold increase in the CAR when moving from Fewer Open to More Open Workplaces. Implementing both maximum work-from-home and cohorts in school along with social distancing measures will reduce transmission, hospitalizations, and deaths. We observe significant heterogeneity within the U.S. due to the starting initial conditions of current cases, local demographic drivers (i.e., age distribution), and the number and type of workplaces. Areas with high incidence at the beginning of the simulation have worse outcomes and, generally, earlier peaks. This could mean that timing school reopening to coincide with locally lower incidence rates is important. Allowing 100% of students to return back to school is likely to lead to additional increases of infection under the current transmission dynamics in the U.S. and if schools reopen at the 80% or 100% level, school-age children could generate the largest number of cases. All scenarios where schools open even at the 80% levels will result in greater COVID-19 case rates requiring higher levels of hospitalizations, ICU beds, and ventilators needed across the U.S. However, implementing cohorts and smaller class sizes result in fewer cases and deaths, while providing important educational opportunities for children. Combining these with social distancing measures including mask wearing, meeting outside, and keeping distanced from others results in many fewer cases.

Our findings should be considered in context of several potential limitations. First, the model assumed the same level of workplace restrictions (namely Fewer Open Workplaces and More Open Workplaces) uniformly across all the states in the United States in order to compare the different scenarios under similar conditions. However, there is evidence that each state has implemented different public health actions resulting in drastically distinct operating statuses for businesses that have reopened [36]. Therefore, incorporating the heterogeneity in state actions may be necessary in order to better quantify the impact of school reopening scenarios on COVID-19 spread. Second, we did not consider testing and contact tracing explicitly in the simulation. Although we assume isolation of symptomatic individuals promptly after symptom onset, we know that effective contact tracing and testing, in combination with hybrid school reopening scenarios and social distancing measures, will be critical for safely reopening schools. Third, we projected epidemic trajectories through the beginning of April 2021 in order to assess the potential impact of school reopening scenarios during the autumn and winter months. However, several studies [37] have shown that behavioral responses to an epidemic or pandemic are highly dependent on the perception of the severity of the disease. Thus, we expect the behavior and compliance to change and fluctuate in the next six months as a result of new public health orders and disease perception; however, we do not have adequate data to predict this, and therefore assume that the same level of restriction and compliance to non-pharmaceutical interventions will remain in place. Fourth, the population distribution in EpiCast is based upon the 2000 U.S. Census data, to take advantage of the tract-to-tract work flow data that was last compiled then. This is a major limitation for areas that have seen significant population changes in the last two decades, therefore, the simulation may be conservative in terms of the potential contacts and spread of COVID-19. Finally, the epidemiological parameters have spatial and temporal variability during the course of the COVID-19 pandemic. Therefore, additional studies are needed in order to quantify the impact of changing these assumptions on the epidemic projections.

While there is uncertainty in our epidemic projections, our results are consistent with previously published studies [18, 26-28] and are intended to serve as guideposts for deliberations regarding the potential relative impact of different school reopening scenarios in the U.S.

## Data Availability

Data are available in the main text or supplementary materials.

## Acknowledgments

The authors wish to acknowledge the following collaborators from the Centers for Disease Control and Prevention (CDC): Dr. Matthew Biggerstaff (Influenza Division), Dr. Rachel Slayton (Division of Healthcare Quality Promotion), and Dr. Amra Uzicanin (Division of Global Migration and Quarantine at CDC). The authors thank Dr. Jee Hwang, economist at the New Mexico Human Services Department for providing useful information to inform workforce assumptions. Additionally, the authors thank the High-Performance Computing team at LANL for their help with the computational infrastructure and for allocating targeted resources to enable COVID-19 modeling efforts.

## Appendix –Figures & Tables

Tables A-1 and A-2 show the key results from the modeling for the Chicago MSA region. Table A-1 shows the total number of cases, deaths, and hospitalizations for each scenario for the full simulation period and for the four weeks around the peak of the epidemic. Table A-2 shows the cumulative and peak incidence (and prevalence) as well as the time to peak and the total CAR for each scenario. It is worth noting that both the 2-day and alternating week school scenarios lead to similar CARs in spite of differences in the reduced social distancing assumptions. For Fewer Open Workplaces, ideal social distancing results in a CAR of 5.8% and 6.5%, respectively for the 2-day and alt-week scenarios, rising to 8.8% and 9.7%, respectively with less social distancing. More Open Workplaces results in significantly higher CAR of 29.2% and 29.7%, respectively with ideal social distancing, and 29.7% and 30.3%, respectively with less social distancing. Likewise, peak timing is spread out significantly under Fewer Open Workplaces due to the impact of these partial reopening scenarios in flattening the curve.

**Table A-1.** Summary of key EpiCast results for Chicago MSA region – Part 1

| Workplace Assumptions | Scenario Name | During Peak 4 Weeks |  |  | August 15, 2020 to April 11, 2021 |  |  |
| --- | --- | --- | --- | --- | --- | --- | --- |
|  |  | Cases | Hospitalized | Deaths | Cases | Hospitalized | Deaths |
| Fewer Open Workplaces | Pre-Pandemic Behavior | 2,159,213 | 60,974 | 3,163 | 3,400,697 | 99,928 | 6,449 |
|  | Baseline | 995,674 | 26,834 | 1,402 | 2,466,645 | 67,484 | 3,867 |
|  | 80%_OL_SD | 527,005 | 14,501 | 787 | 1,876,690 | 52,036 | 2,980 |
|  | 80%_OL_LessSD | 660,681 | 18,117 | 965 | 2,068,149 | 57,186 | 3,253 |
|  | 40%_POL_SD_Week | 82,602 | 2,348 | 138 | 515,501 | 14,401 | 818 |
|  | 40%_POL_SD_2Days | 70,226 | 1,990 | 117 | 457,062 | 12,810 | 733 |
|  | 40%_POL_LessSD_Week | 134,122 | 3,773 | 220 | 763,588 | 21,187 | 1,207 |
|  | 40%_POL_LessSD_2Days | 119,614 | 3,392 | 185 | 694,637 | 19,392 | 1,084 |
|  | Offsite | 45,415 | 1,286 | 76 | 339,314 | 9,571 | 558 |
| More Open Workplaces | Pre-Pandemic Behavior | 2,387,805 | 69,185 | 3,669 | 3,586,058 | 106,262 | 6,804 |
|  | Baseline | 1,811,617 | 50,808 | 2,587 | 3,206,736 | 92,648 | 5,679 |
|  | 80%_OL_SD | 1,397,360 | 40,040 | 2,200 | 2,928,719 | 85,311 | 5,293 |
|  | 80%_OL_LessSD | 1,450,481 | 41,658 | 2,274 | 2,964,029 | 86,382 | 5,316 |
|  | 40%_POL_SD_Week | 712,232 | 21,100 | 1,270 | 2,337,759 | 69,556 | 4,361 |
|  | 40%_POL_SD_2Days | 682,950 | 20,261 | 1,222 | 2,297,503 | 68,493 | 4,282 |
|  | 40%_POL_LessSD_Week | 747,026 | 22,083 | 1,336 | 2,382,261 | 70,857 | 4,469 |
|  | 40%_POL_LessSD_2Days | 717,028 | 21,208 | 1,275 | 2,341,499 | 69,694 | 4,374 |
|  | Offsite | 579,268 | 17,320 | 1,055 | 2,107,852 | 63,272 | 3,976 |

**Table A-2.** Summary of key EpiCast results for the Chicago MSA region – Part 2

| Workplace Assumptions | Scenario Name | Peak Incidence (Cases per 1000) | Time to Peak Incidence (days) | Peak Prevalence (Cases per 1000) | Time to Peak Prevalence (days) | Clinical Attack Rate (%) |
| --- | --- | --- | --- | --- | --- | --- |
| **Fewer Open Workplaces** | **Pre-Pandemic Behavior** | 92,048 | 69 | 63.4 | 72 | 43.2 |
|  | **Baseline** | 38,677 | 97 | 26.9 | 101 | 31.3 |
|  | **80%_OL_SD** | 19,844 | 125 | 13.9 | 129 | 23.8 |
|  | **80%_OL_LessSD** | 24,911 | 118 | 17.4 | 122 | 26.3 |
|  | **40%_POL_SD_Week** | 3,011 | 188 | 2.1 | 205 | 6.5 |
|  | **40%_POL_SD_2Days** | 2,559 | 201 | 1.8 | 206 | 5.8 |
|  | **40%_POL_LessSD_Week** | 4,911 | 188 | 3.5 | 192 | 9.7 |
|  | **40%_POL_LessSD_2Days** | 4,383 | 188 | 3.1 | 192 | 8.8 |
|  | **Offsite** | 1,648 | 174 | 1.2 | 178 | 4.3 |
| **More Open Workplaces** | **Pre-Pandemic Behavior** | 102,256 | 65 | 71.4 | 70 | 45.6 |
|  | **Baseline** | 74,889 | 76 | 51.8 | 80 | 40.7 |
|  | **80%_OL_SD** | 54,907 | 83 | 38.5 | 87 | 37.2 |
|  | **80%_OL_LessSD** | 57,771 | 83 | 40.3 | 87 | 37.6 |
|  | **40%_POL_SD_Week** | 26,489 | 110 | 18.7 | 115 | 29.7 |
|  | **40%_POL_SD_2Days** | 25,471 | 117 | 17.9 | 119 | 29.2 |
|  | **40%_POL_LessSD_Week** | 27,952 | 110 | 19.7 | 115 | 30.3 |
|  | **40%_POL_LessSD_2Days** | 26,823 | 110 | 18.9 | 115 | 29.7 |
|  | **Offsite** | 21,484 | 118 | 15.2 | 122 | 26.8 |

Figures B-1 to B-7 show epidemic time series for 12 states for all the scenarios not shown in the main manuscript for both Fewer Open and More Open Workplaces. Data for all other states (including the ones shown here) are provided as supplemental material under Supplemental Files SF1-SF3.

**Figure B-1.**
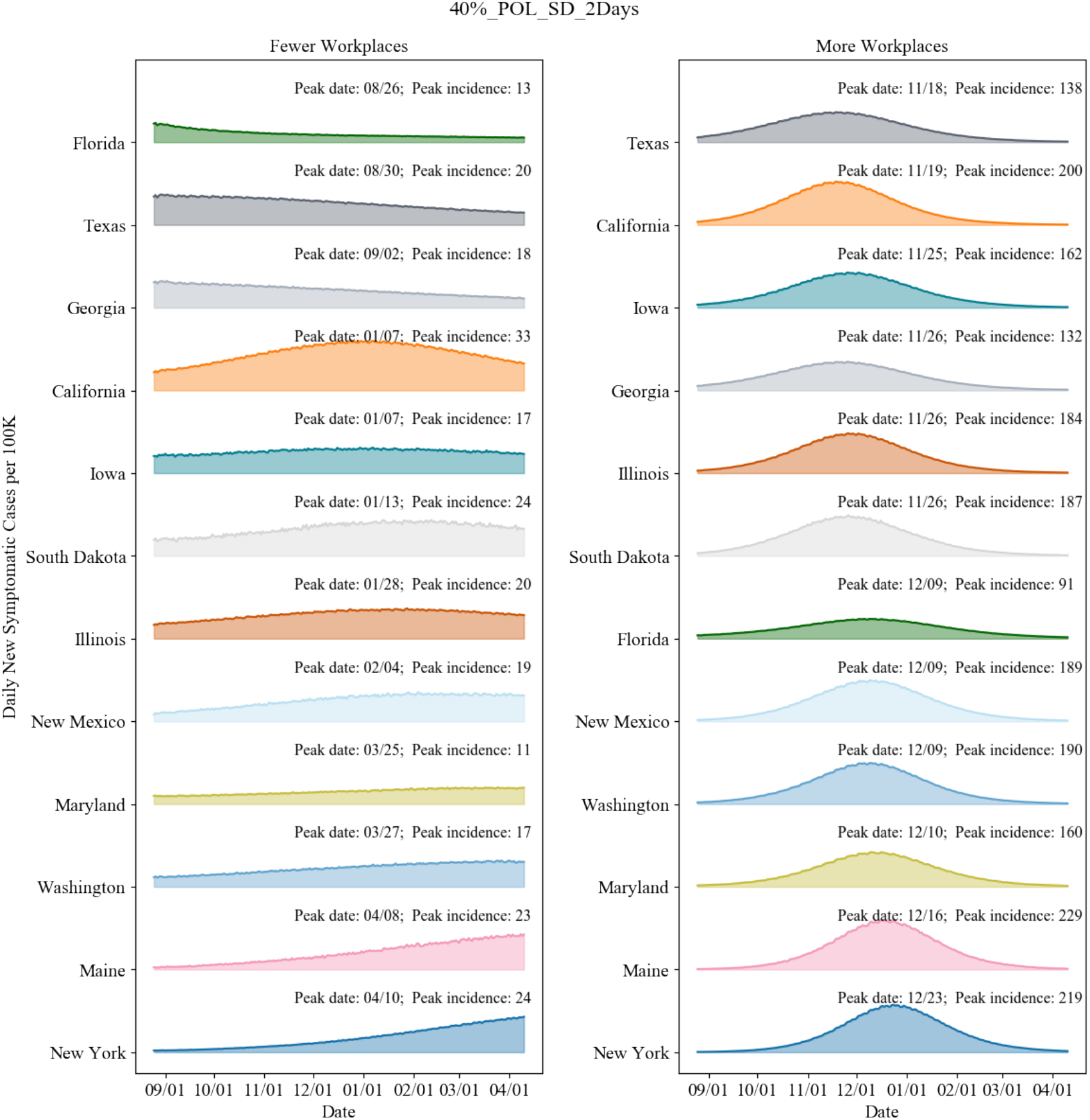
40% onsite 2-day epidemic curves and peak dates for 12 representative states. (States are sorted by peak date for both scenarios.)

**Figure B-2.**
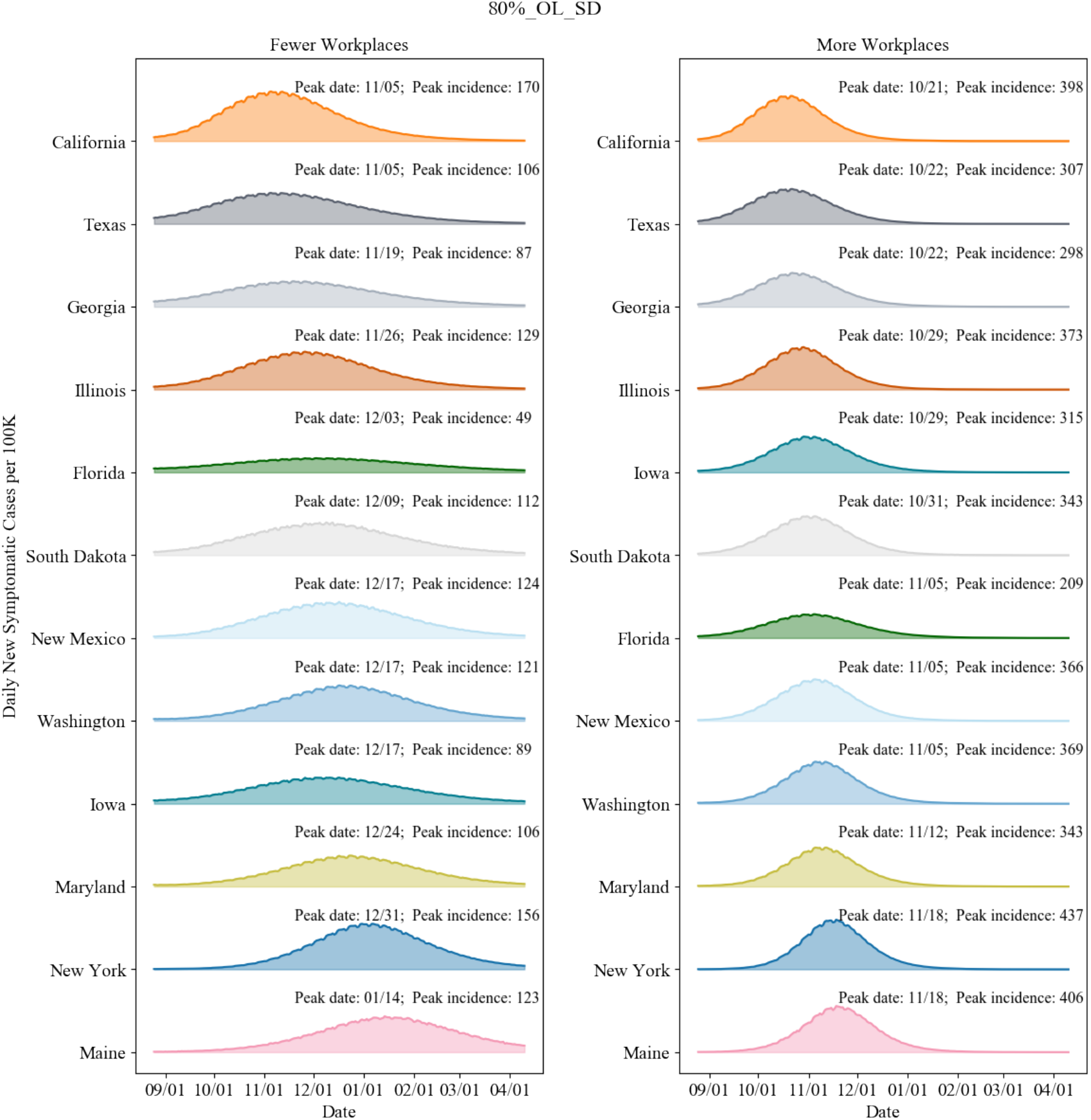
80% onsite epidemic curves and peak dates for 12 representative states. (States are sorted by peak date for both scenarios.)

**Figure B-3.**
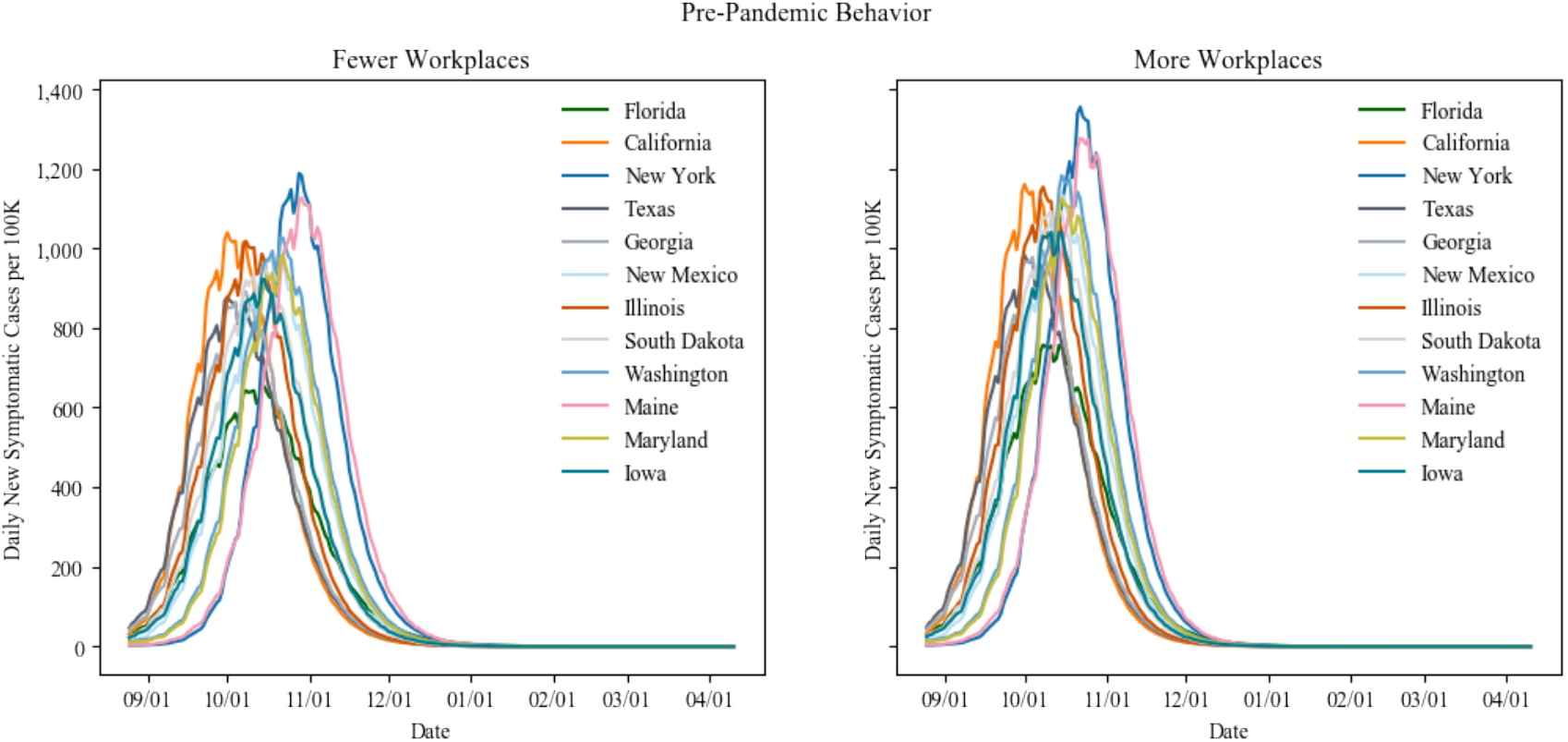
Pre-pandemic Behavior epidemic curves for 12 representative states

**Figure B-4.**
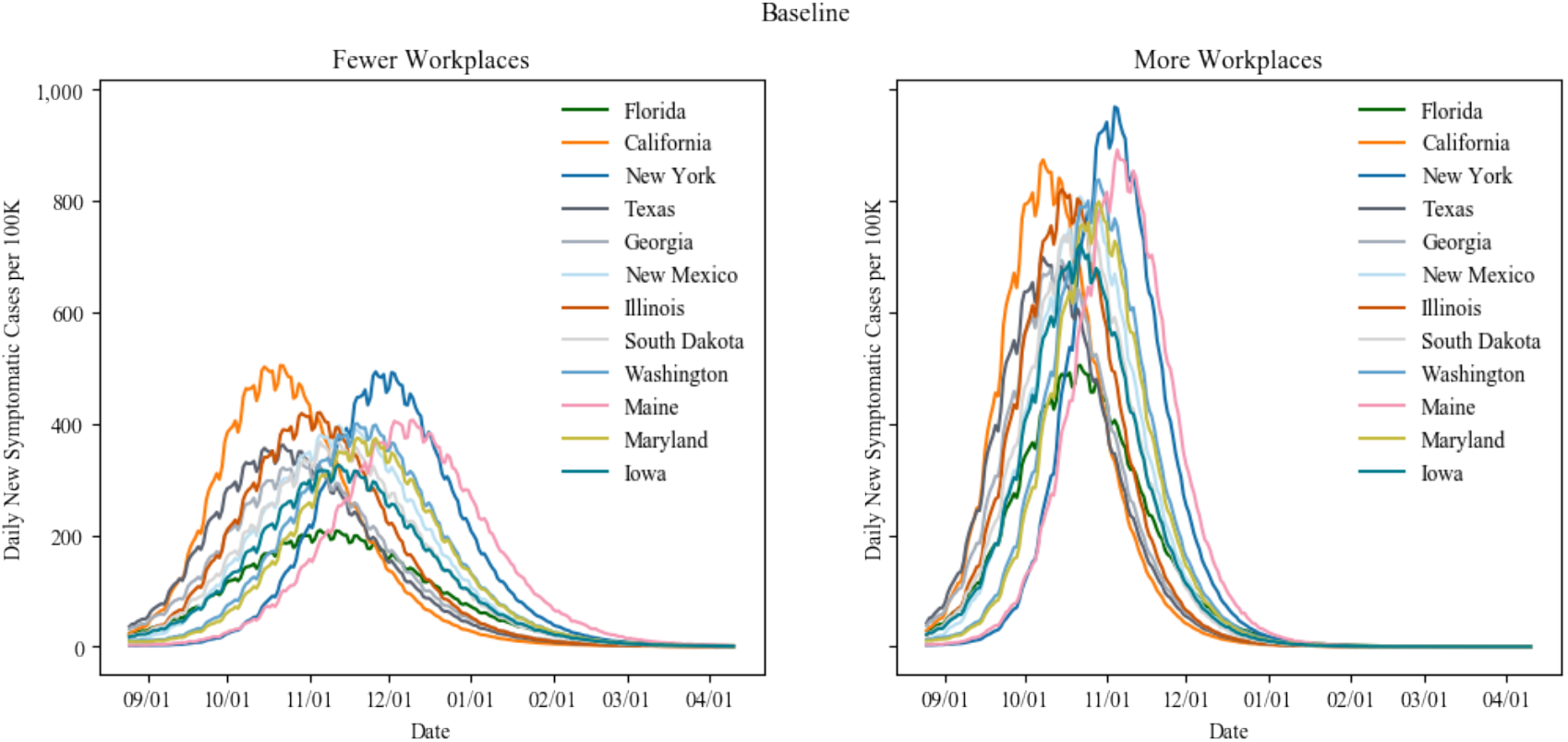
Baseline epidemic curves for 12 representative states

**Figure B-5.**
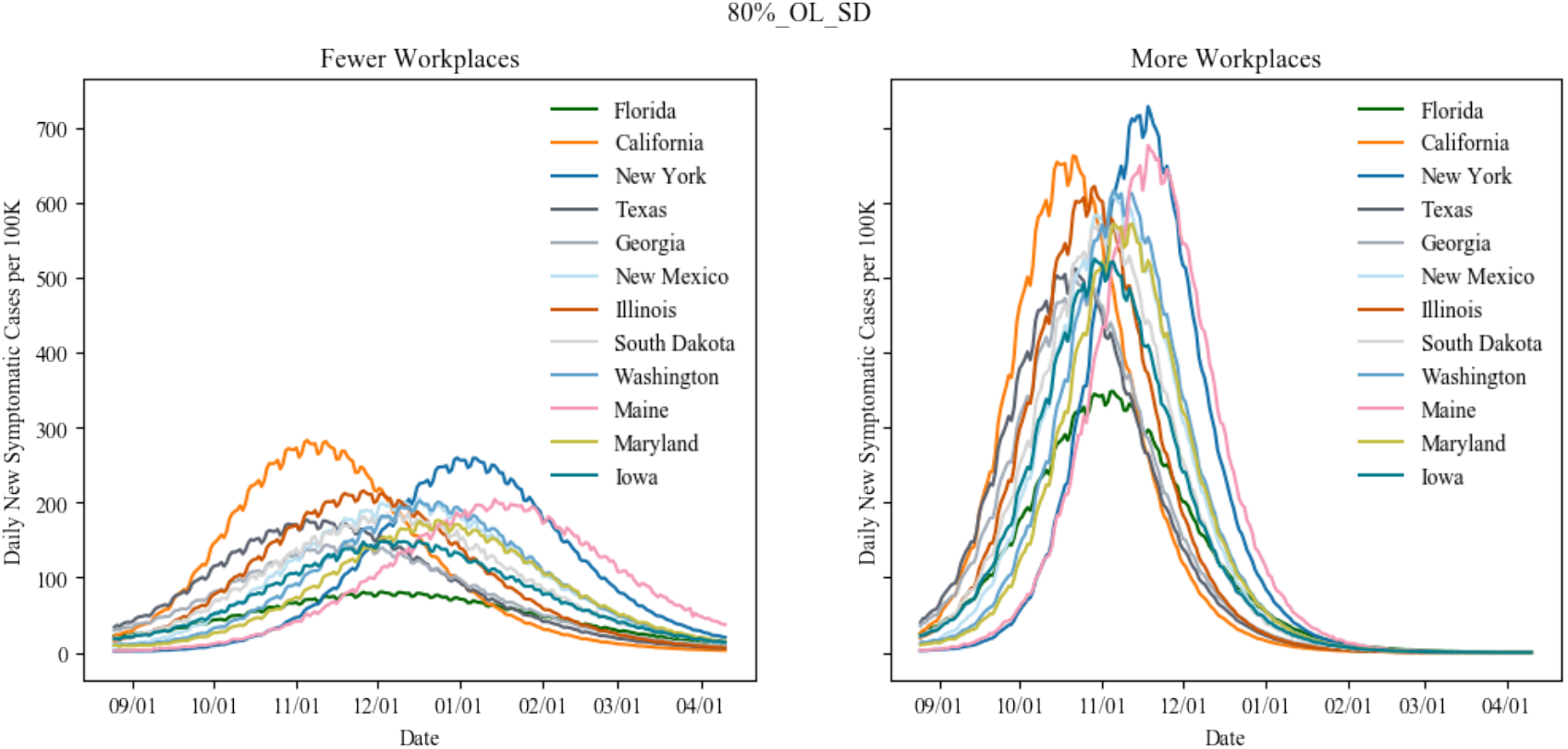
80% onsite epidemic curves for 12 representative states

**Figure B-6.**
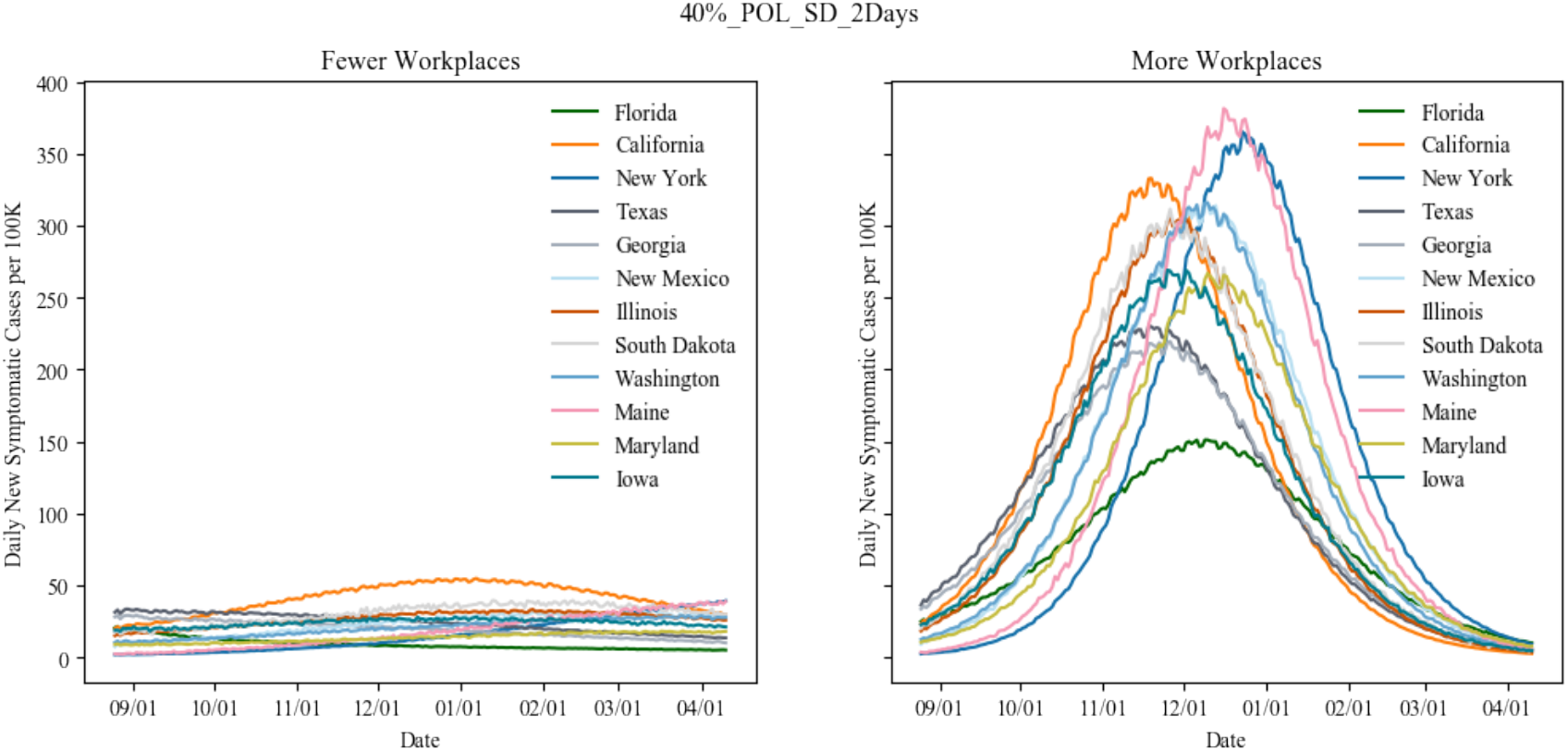
40% onsite 2-day epidemic curves for 12 representative states

**Figure B-7.**
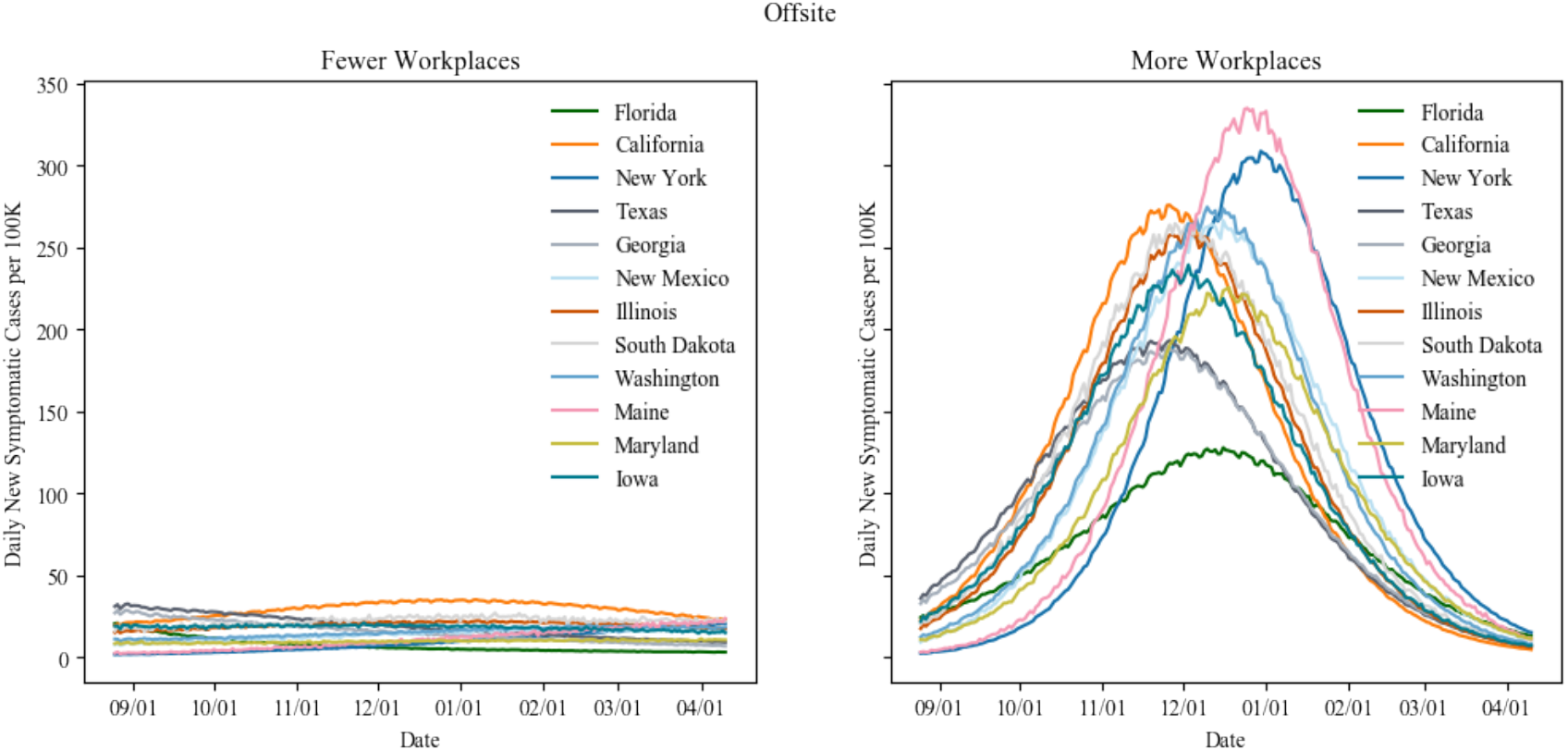
Offsite epidemic curves for 12 representative states

Figure B-8 shows the source of infection as a percentage aggregated at the national-level for all the scenarios for both Fewer Open and More Open Workplaces.

**Figure B-8.**
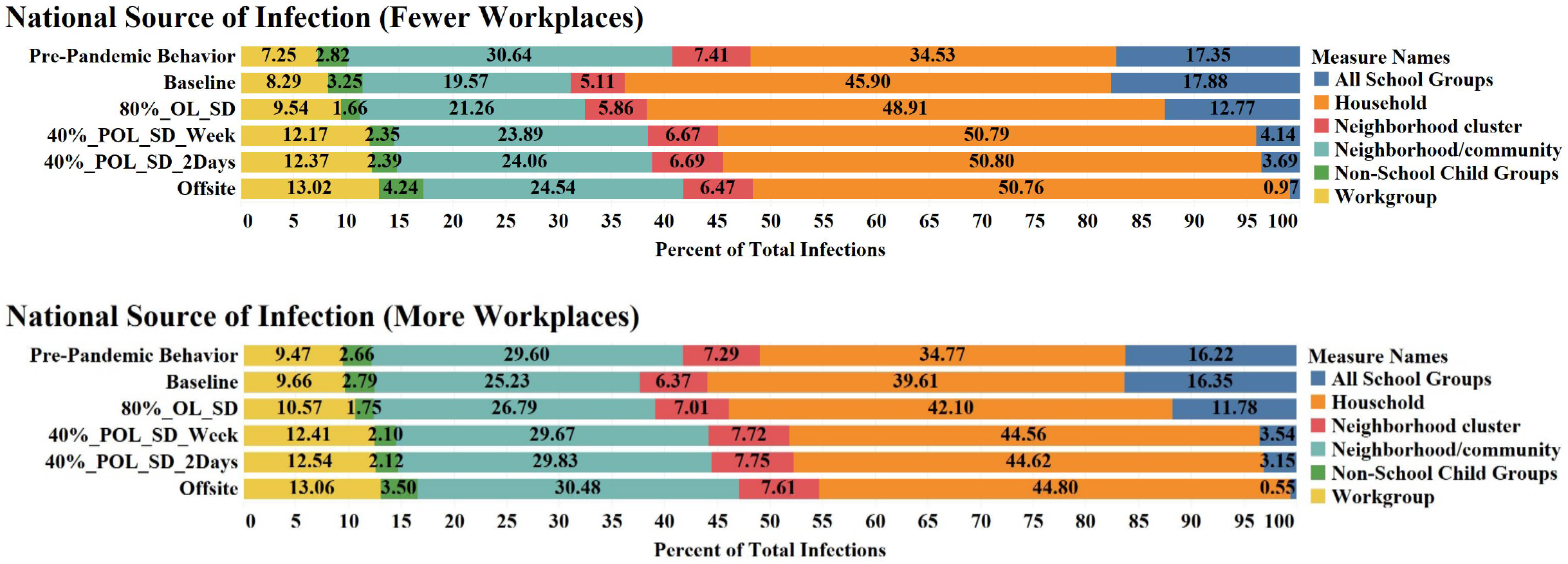
Source of infection for scenarios aggregated for all states.

Figure B-9 shows the total number of cases, hospitalizations, and ICU beds by age group for all the national-level scenarios for More Open Workplaces.

**Figure B-9.**
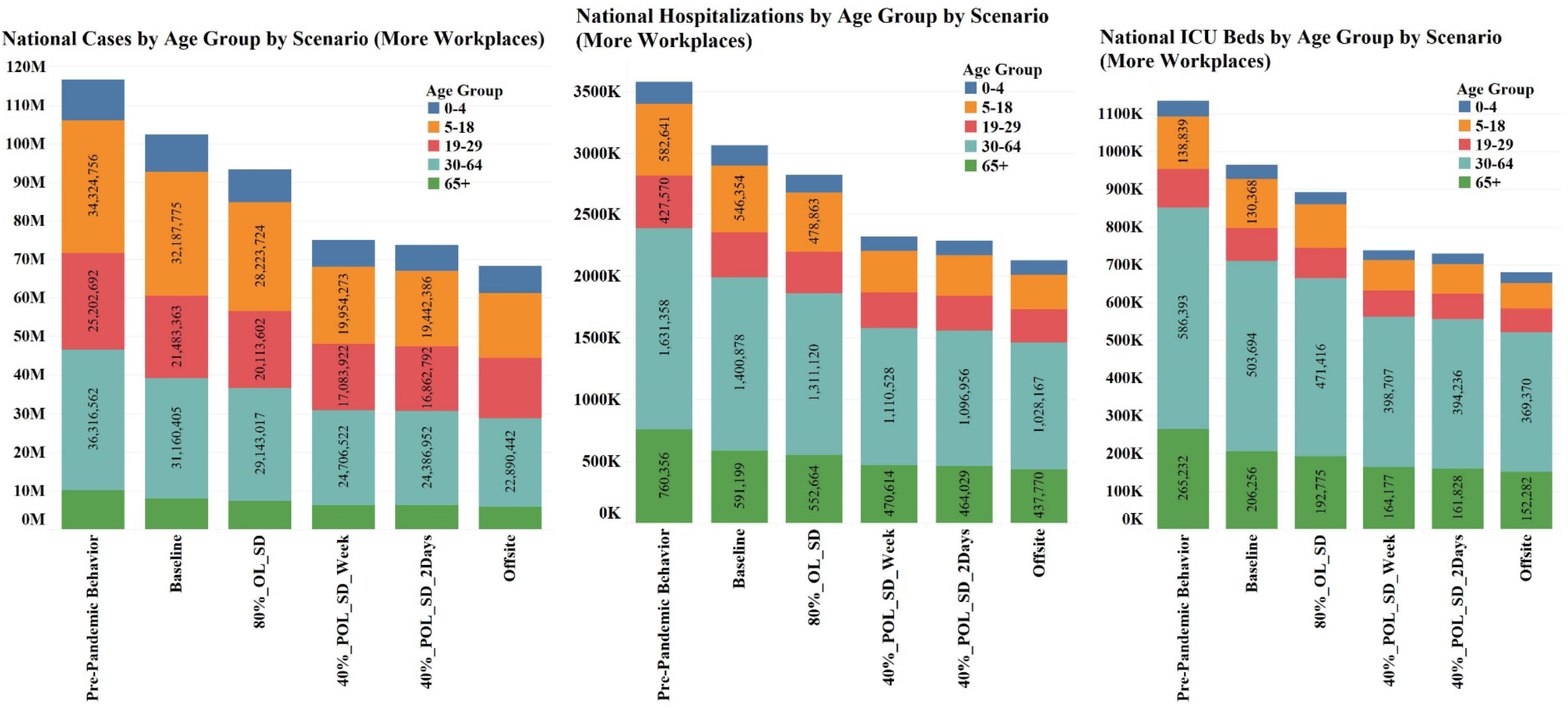
Total cases, hospitalizations, and ICU beds by age group for all the national-level scenarios for More Open Workplaces.

**Figure B-10.**
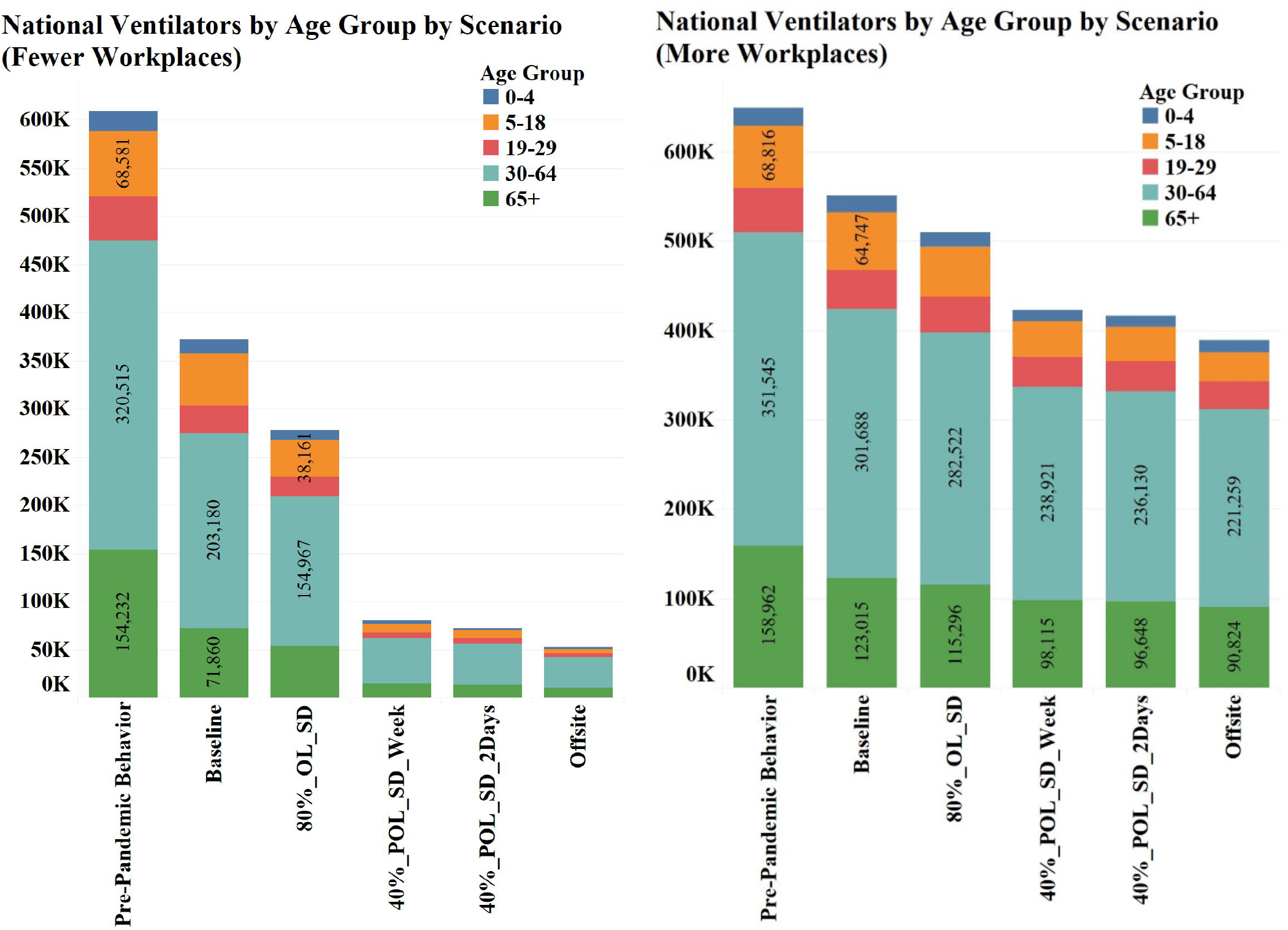
Total ventilators used by age group for all national-level scenarios.

## Notes

### Competing Interest Statement

The authors have declared no competing interest.

### Clinical Trial

N/A

### Funding Statement

This work was funded by the United States Centers for Disease Control and Prevention.

### Author Declarations

This is a purely modeling and simulation study. While we used publicly available COVID-19 cases from the NYT GitHub repository to initialize our model, the research involves no intervention or interaction [102(f)(1) and (2)], and the data are publicly available [102(f)(2)]. Hence, our paper is not human subjects research veered by 45 CFR part 46.

